# A phase 2a double-blind, placebo-controlled randomized trial of the SARS-CoV-2-specific monoclonal antibody AER002 in people with Long COVID

**DOI:** 10.64898/2026.03.07.26347857

**Authors:** Michael J. Peluso, Dylan Ryder, Thomas Dalhuisen, Danny Hoi Tsun Chu, Meghann C. Williams, Antonio E. Rodriguez, Brian LaFranchi, Joanna Vinden, Emily A. Fehrman, Beatrice Huang, Rebecca Hoh, Kofi A. Asare, Kathleen Bellon Pizarro, Mohammad Rahman, Emilio de Narvaez, Mark M. Painter, E. John Wherry, Zoe N. Swank, Louise L. Hansen, David R. Walt, Yoshinori Fukazawa, Anisha Sekar, Steven E. Bellan, Holly Tieu, Josephat Asiago, Prakash Bhuyan, Rajeev Venkayya, Robert R. Flavell, Henry VanBrocklin, J. Daniel Kelly, Priscilla Y. Hsue, Matthew S. Durstenfeld, Peter W. Hunt, Leonard Calabrese, Ma Somsouk, Jeffrey N. Martin, David V. Glidden, Amelia N. Deitchman, Timothy J. Henrich, Steven G. Deeks

**Affiliations:** Division of HIV, Infectious Diseases, and Global Medicine, University of California, San Francisco, San Francisco, CA, USA; Department of Clinical Pharmacy, University of California, San Francisco, San Francisco, CA, USA; Division of Experimental Medicine, University of California, San Francisco, San Francisco, CA, USA; Division of Cardiology, University of California, San Francisco, San Francisco, CA, USA; University of Pennsylvania, Philadelphia, PA, USA; Department of Pathology, Brigham & Women’s Hospital, Harvard Medical School, Boston, MA, USA; Patient-Led Research Collaborative; Aerium Therapeutics, Boston, MA, USA; Department of Radiology and Biomedical Imaging, University of California, San Francisco, San Francisco, CA, USA; Department of Epidemiology and Biostatistics, University of California, San Francisco, San Francisco, CA, USA; Division of Cardiology, University of California, Los Angeles, CA, USA; Cleveland Clinic, Cleveland, OH, USA; Division of Gastroenterology, University of California, San Francisco, San Francisco, CA, USA

**Keywords:** Long COVID, post-acute sequelae of SARS-CoV-2, viral persistence, monoclonal antibody

## Abstract

Long COVID is a disabling chronic illness with no proven treatments. Persistence of SARS-CoV-2 has been proposed as a biological driver of the disease. We conducted a placebo-controlled, double-blind, 2:1 randomized mechanistic trial of the SARS-CoV-2-specific monoclonal antibody AER002 in 36 participants who met the World Health Organization case definition of Long COVID. After baseline characterization, participants received a single infusion and were followed for 360 days. The primary endpoint was the PROMIS-29 Physical Health Summary Score (PHSS) at 90 days; secondary and exploratory endpoints included patient-reported and objective measures of physical, cognitive, and neurologic function as well as blood-, imaging-, and tissue-based biomarkers. While AER002 was safe and well tolerated, no significant differences in physical health, quality of life, objective measures of physical function or cognition, or blood-based biomarkers were demonstrated between the treatment and control arms. In a post-hoc analysis, participants with a lower baseline SARS-CoV-2 antibody level and higher drug exposure were more likely to express a perceived treatment benefit based on the Patient Global Impression of Change scale (p<0.05 for anti-S, S1, and RBD). Although AER002 was not efficacious in this proof-of-concept study of people with broadly defined Long COVID, our findings could inform recruitment or dosing strategies employed in future trials using monoclonal antibodies to target viral persistence as a driver of Long COVID.

## BACKGROUND

Long COVID is a debilitating infection-associated chronic condition (IACC) characterized by COVID-attributed symptoms that persist for months to years following a SARS-CoV-2 infection^1^. Millions of individuals are affected worldwide^2^ and the health and economic impacts are profound^3^. Despite its public health significance, the pathobiology that drives Long COVID is incompletely understood^4^. There is thus an urgent need for rigorous, randomized trials not only to identify promising therapeutics but also to shed light on the underlying pathobiological processes that drive the condition^5^.

Although SARS-CoV-2 infection was initially assumed to be transient, there is growing evidence to suggest that components of the virus (RNA and/or protein) can persist for months or years in some individuals^6^. Multiple studies have now demonstrated SARS-CoV-2 antigens circulating in blood for months after acute infection^7–9^, although these findings are not universal^10^. Others have identified SARS-CoV-2 RNA or protein in tissue sites, including in tissue acquired via gut mucosal biopsies^11–16^. The nature of this persistence – whether it represents inert RNA, RNA that can be translated to produce viral proteins, or even low-level viral replication – remains unclear^17^. Also unclear is whether SARS-CoV-2 persistence is a direct cause of Long COVID or a consequence of another process (e.g., immune dysregulation) that is the primary driver of the condition^5^.

In response to case reports suggesting a benefit of off-label antiviral use in at least some people with Long COVID^18–20^, several clinical trials of antivirals have been undertaken. Two trials testing a 15-day course of the protease inhibitor nirmatrelvir/ritonavir (brand name: Paxlovid) failed to show significant benefit in improving symptoms and quality of life among individuals with Long COVID^21,22^. However, this strategy has several limitations, the most notable of which are the short duration of treatment and that drugs like nirmatrelvir would only be expected to have an effect if Long COVID were driven by active replication of SARS-CoV-2^5,17^.

SARS-CoV-2-specific monoclonal antibodies have been proposed as another strategy to address viral persistence^4,5,17,23^. Initially developed for treatment or prevention of SARS-CoV-2 infection, these drugs work by binding and neutralizing virions and preventing infection of new target cells^24^. In some cases, monoclonal antibodies can be modified to have an extended half-life^25–27^, allowing for a prolonged therapeutic or prophylactic effect. Furthermore, in cases in which effector function is maintained, such antibodies also may theoretically facilitate opsonization of virions and/or clearance of infected cells expressing spike protein via antibody- or complement-mediated cellular cytotoxicity, even in the absence of neutralization^28^. Several case series have detailed profound improvement in Long COVID symptoms after a single infusion of SARS-CoV-2-specific monoclonal antibodies in a small number of individuals^29,30^. Despite this anecdotal evidence, there are no published randomized clinical trials using monoclonal antibodies for Long COVID.

Here, we report the results of outSMART-LC (NCT05877508), a 2:1 randomized, double-blind, placebo-controlled phase 2a trial of a single infusion of the long-acting SARS-CoV-2-specific monoclonal antibody AER002^31^ in people with Long COVID. AER002 is a fully human monoclonal immunoglobulin-G1 (IgG1) mAb that binds to the receptor binding domain (RBD) of SARS-CoV-2 spike protein^31^. The antibody demonstrated in vitro activity against SARS-CoV-2 variants of concern including Omicron strains BA.2.12.1, BA.4., and BA.5. AER002 also retains some activity against the BQ.1 variant, albeit at a lower level than earlier Omicron variants. AER002 included modifications to the fragment crystallizable (Fc) part of the antibody to prolong half-life. Despite its waning efficacy against later SARS-CoV-2 variants, we theorized AER002 would remain effective in clearing SARS-CoV-2 persistence from infections occurring before it lost neutralizing capacity.

We demonstrate that while AER002 was safe and well tolerated in people with Long COVID, it was ineffective in reducing Long COVID symptoms, enhancing quality of life, or improving objective performance measures compared to placebo in this broadly defined population. However, exploratory analyses suggested a biological effect on tissue-based immune activation and revealed that higher drug exposure was associated with clinical response, informing the design of future monoclonal antibody trials targeting viral persistence in Long COVID.

## RESULTS

### Cohort description

From July 2023 to April 2024, a total of 460 individuals expressed interest in the trial. Forty-three consented to participation and underwent eligibility assessment and 36 were enrolled and randomized to receive AER002 versus placebo in a 2:1 ratio (Figure 1). Median age was 42 years (interquartile range [IQR]: 32-53); 56% of participants were female sex; 75% were White, 14% Asian, 8% Hispanic/Latino, and 3% Black (Table 1). Pre-existing medical comorbidities included: lung problems (asthma, COPD, emphysema, or bronchitis; 17%), autoimmune disease (14%), hypertension (11%), diabetes (2.8%), and HIV (2.8%). Participants with a diagnosis of myalgic encephalomyelitis/chronic fatigue syndrome (ME/CFS) preceding their SARS-CoV-2 infection were excluded. Prior to infusion, 5.6% of participants had received a mast cell activation syndrome (MCAS) diagnosis and 8.3% met criteria for hypermobile Ehlers-Danlos Syndrome. Participants of female sex were also evaluated for pain symptoms related to endometriosis (5.6% score ≥60).

**Figure 1.**
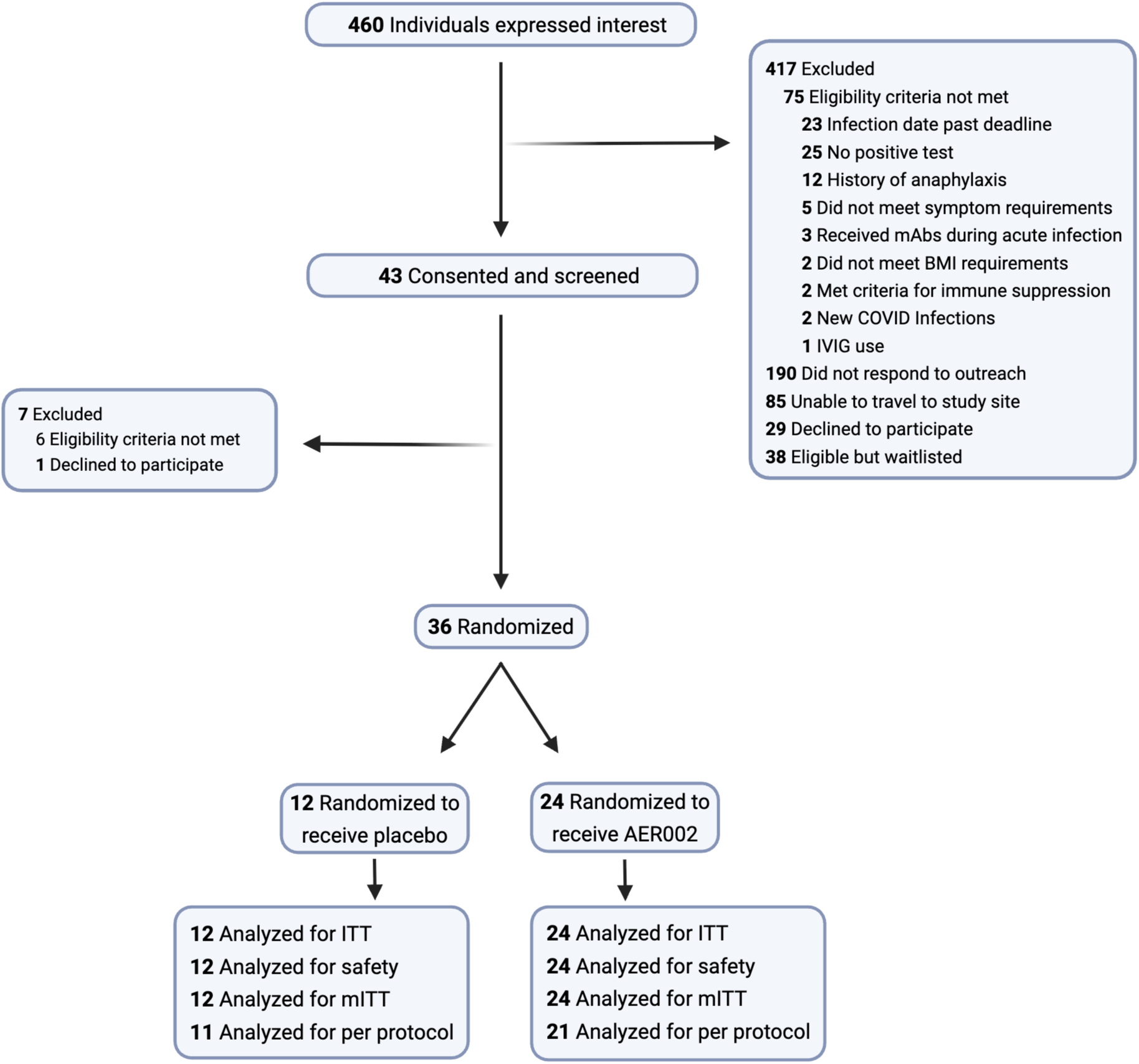
CONSORT diagram for the clinical trial.

**Table 1.** Demographics of the study cohort.

| Characteristic | Overall<br>N = 36<sup>1</sup> | AER002<br>N = 24<sup>1</sup> | Placebo<br>N = 12<sup>1</sup> |
| --- | --- | --- | --- |
| **Age at enrollment (years)** | 42 (32-53) | 42 (33-49) | 43 (30-59) |
| **Sex at birth** |  |  |  |
| Female | 20 (56%) | 13 (54%) | 7 (58%) |
| Male | 16 (44%) | 11 (46%) | 5 (42%) |
| **Gender** |  |  |  |
| Female | 19 (53%) | 12 (50%) | 7 (58%) |
| Male | 16 (44%) | 11 (46%) | 5 (42%) |
| Non-binary | 1 (2.8%) | 1 (4.2%) | 0 (0%) |
| **Race/Ethnicity** |  |  |  |
| Asian | 5 (14%) | 3 (13%) | 2 (17%) |
| Black/African American | 1 (2.8%) | 0 (0%) | 1 (8.3%) |
| Hispanic/Latino | 3 (8.3%) | 2 (8.3%) | 1 (8.3%) |
| White | 27 (75%) | 19 (79%) | 8 (67%) |
| **Medical comorbidities** |  |  |  |
| Autoimmune disease | 5 (14%) | 2 (8.3%) | 3 (25%) |
| Cancer<sup>2</sup> | 0 (0%) | 0 (0%) | 0 (0%) |
| Diabetes | 1 (2.8%) | 1 (4.2%) | 0 (0%) |
| Heart attack or heart failure | 0 (0%) | 0 (0%) | 0 (0%) |
| HIV Infection | 1 (2.8%) | 0 (0%) | 1 (8.3%) |
| Hypertension | 4 (11%) | 3 (13%) | 1 (8.3%) |
| Lung problems<sup>3</sup> | 6 (17%) | 4 (17%) | 2 (17%) |
| Kidney disease | 0 (0%) | 0 (0%) | 0 (0%) |
| **Body mass index (kg/m<sup>2</sup>)** |  |  |  |
| 18.0-18.5 | 1 (2.8%) | 1 (4.2%) | 0 (0%) |
| 18.5-24.9 | 21 (58%) | 14 (58%) | 7 (58%) |
| 25.0-29.9 | 7 (19%) | 4 (17%) | 3 (25%) |
| >=30 | 7 (19%) | 5 (21%) | 2 (17%) |
| **ME/CFS diagnosis preceding SARS-CoV-2 infection** | 0 (0%) | 0 (0%) | 0 (0%) |
| **Mast Cell Activation Syndrome** | 2 (5.6%) | 0 (0%) | 2 (17%) |
| **Beighton score<sup>4</sup>** |  |  |  |
| 0 | 27 (75%) | 21 (88%) | 6 (50%) |
| 2-4.9 | 6 (17%) | 1 (4.2%) | 5 (41.7%) |
| >=5 | 3 (8.3%) | 2 (8.3%) | 1 (8.3%) |
| **ENDOPAIN-4D global score<sup>5</sup>** |  |  |  |
| 0 | 12 (33%) | 6 (25%) | 6 (50%) |
| 0.1-29.9 | 4 (11.1%) | 3 (13%) | 1 (8.3%) |
| 30-59.9 | 2 (5.6%) | 2 (8.3%) | 0 (0%) |
| >=60 | 2 (5.6%) | 2 (8.3%) | 0 (0%) |

<sup>1</sup> n (%) Median (IQR)<sup>2</sup> Cancer requiring treatment within the 2 years before COVID-19<sup>3</sup> Asthma, COPD, emphysema, or bronchitis experienced in the 5 years before COVID-19<sup>4</sup> A score of 5+ indicates a positive indication for Hypermobility Ehlers-Danlos Syndrome<sup>5</sup> Among those with female sex at birth

Most participants (53%) reported a single SARS-CoV-2 infection prior to the study (Table 2); the SARS-CoV-2 infection to which Long COVID symptoms were attributed occurred a median of 740 days (IQR: 576-1113) prior to enrollment; few participants (5.6%) had been previously hospitalized for COVID-19. The most recent infection was a median of 504 days (IQR 344-706) prior to enrollment. Notably 33% of participants had experienced a SARS-CoV-2 reinfection after August 15, 2022, when AER002 was expected to have lost neutralizing capacity. All participants had received at least one SARS-CoV-2 vaccine prior to participation in the study; in 31% of cases, the initial SARS-CoV-2 infection had preceded vaccination. The cohort was highly symptomatic. In the 30 days preceding enrollment, participants reported a median of 14 Long COVID symptoms (IQR: 7.5-18.5). The most common symptoms were constitutional (e.g., fatigue; 92%), neurologic (e.g., “brain fog”, headache; 100%), and cardiopulmonary (e.g., exercise intolerance; 81%); 31 (86%) participants reported post-exertional malaise (PEM; “Symptoms that get worse more frequently than usual after physical or mental effort, such as fatigue, pain, or swollen glands”) at baseline and 21 (58%) met the 2015 Institute of Medicine criteria for ME/CFS^32^. Both groups reported significant impact of Long COVID symptoms on functional status and quality of life. At baseline, the median quality of life on a 100-point visual analogue scale was 49 (IQR 35-63).

**Table 2.** COVID-19 and Long COVID history of the study cohort.

| Characteristic | Overall<br>N = 36<sup>1</sup> | AER002<br>N = 24<sup>1</sup> | Placebo<br>N = 12<sup>1</sup> |
| --- | --- | --- | --- |
| **Number of SARS-CoV-2 infections at time of enrollment** |  |  |  |
| 1 | 19 (53%) | 13 (54%) | 6 (50%) |
| 2 | 14 (39%) | 9 (38%) | 5 (42%) |
| 3 | 3 (8.3%) | 2 (8.3%) | 1 (8.3%) |
| **Index SARS-CoV-2 infection date** |  |  |  |
| March 2020 - April 2021 | 12 (33%) | 8 (33%) | 4 (33%) |
| May 2021 - November 2021 | 3 (8.3%) | 2 (8.3%) | 1 (8.3%) |
| December 2021 - August 2022 | 21 (58%) | 14 (58%) | 7 (58%) |
| **Time since infection to which Long COVID is attributed (days)** | 740 (576-1113) | 740 (571-1113) | 769 (593-1114) |
| **Hospitalized for COVID-19** | 2 (5.6%) | 2 (8.3%) | 0 (0%) |
| **Time since most recent SARS-CoV-2 infection (days)** | 504 (344-706) | 509 (353-709) | 504 (280-662) |
| **Reported a SARS-CoV-2 re-infection after August 15, 2022** | 12 (33%) | 8 (33%) | 4 (33%) |
| **Completed primary SARS-CoV-2 vaccination series** | 36 (100%) | 24 (100%) | 12 (100%) |
| **SARS-CoV-2 infection preceding first SARS-CoV-2 vaccination** | 11 (31%) | 7 (29%) | 4 (33%) |
| **Time since most recent SARS-CoV-2 vaccination (days)** | 422 (138-700) | 456 (190-700) | 327 (99-673) |
| **Long COVID symptom count (last 30 days)** | 14.0 (7.5-18.5) | 12.0 (6.0-16.5) | 14.5 (11.0-20.0) |
| **Symptoms reported (last 30 days)<sup>2</sup>** |  |  |  |
| Constitutional | 33 (92%) | 22 (92%) | 11 (92%) |
| Upper respiratory | 14 (39%) | 9 (38%) | 5 (42%) |
| Cardiopulmonary | 29 (81%) | 19 (79%) | 10 (83%) |
| Gastrointestinal | 22 (61%) | 13 (54%) | 9 (75%) |
| Genitourinary | 6 (17%) | 5 (21%) | 1 (8.3%) |
| Dermatologic | 6 (17%) | 3 (13%) | 3 (25%) |
| Musculoskeletal | 24 (67%) | 16 (67%) | 8 (67%) |
| Neurologic | 36 (100%) | 24 (100%) | 12 (100%) |
| Trouble sleeping | 27 (75%) | 17 (71%) | 10 (83%) |
| Post Exertional Malaise | 31 (86%) | 20 (83%) | 11 (92%) |
| **Met 2015 IOM case definition for ME/CFS<sup>3</sup>** | 21 (58%) | 14 (58%) | 7 (58%) |
| **Quality of Life by Visual Analogue Scale at baseline** | 49 (35-63) | 50 (35-64) | 44 (35-55) |

<sup>1</sup> n (%) Median (IQR)
<sup>2</sup> **Constitutional:** fatigue, fever, chills. **Upper respiratory:** rhinorrhea, sore throat. **Cardiopulmonary:** cough, shortness of breath, heart palpitations, chest pain, fainting spells. **Gastrointestinal:** diarrhea, nausea, loss of appetite, abdominal pain, vomiting, constipation. **Genitourinary:** menstrual cramps, dyspareunia. **Dermatologic:** rash. **Musculoskeletal:** back pain, myalgia, joint pain. **Neurologic:** anosmia, dysgeusia, concentration problems, dizziness, balance problems, neuropathy, vision problems, parosmia.
<sup>3</sup> Participant reports fatigue, post-exertional malaise, and unrefreshing sleep with cognitive impairment and/or dizziness

### Follow-up

All participants completed the trial. All participants remained blinded to treatment allocation until every participant had completed the 180-day timepoint; participants were then followed in an open-label manner for safety until the 360-day timepoint. There were 12 confirmed or suspected COVID-19 reinfections during the study period. Notably, 4 reinfections (3 confirmed, 1 suspected) occurred between the infusion and 90-day timepoints, 3 reinfections (2 confirmed, 1 suspected) occurred between the 90-day and 180-day timepoints, and 5 reinfections (5 confirmed, 0 suspected) occurred between the 180-day and 360-day timepoints.

### Safety and tolerability

AER002 was safe and well-tolerated. A total of 112 adverse events (AEs) among 32 participants were recorded during the study period (Supplemental Tables 1 and 2). The most common adverse events were confirmed acute COVID-19 (9%), COVID-19-negative upper respiratory infection (8%), exacerbation of post-exertional malaise (PEM; 6.3%), and diarrhea (3.6%). One grade 3 AE (a planned hospital admission for management of intractable chronic migraine with aura) occurred during the study but was not related. No clinically significant laboratory abnormalities occurred.

Based on input from people with lived experience of Long COVID and because of the intensity of the study protocol, we paid particular attention to PEM. Using a modified version of the DePaul Symptom Questionnaire for PEM^33^, 29 participants met criteria for PEM prior to the infusion; of those who had baseline PEM, 76% (22/29) reported the presence of PEM in the two days following infusion and 24% (7/29) reported no PEM in the same timeframe. Of 7 without baseline PEM, none reported the emergence of post-infusion PEM in the two days following the infusion. Additionally, we worked to provide accommodations for PEM symptoms during study participation, which ranged from small adjustments during in-person visits (dimmed lights, frequent breaks, access to examination beds for rest) to administration of questionnaires remotely for those too unwell to complete them in person.

### Overall clinical outcomes

#### Physical health and quality of life

In comparison to those receiving placebo, we did not observe a significant improvement in functional status or overall quality of life in those receiving AER002 (Table 3). Among patient-reported outcomes and objective measures, both groups were similar at baseline (Supplemental Table 3). The PROMIS-29 Physical Health Summary Score^34–36^ improved in both groups between baseline and the 90-day timepoint, with no significant difference between groups (Figure 2a). Similarly, the PROMIS-29 Mental Health Summary Score (MHSS) secondary outcome improved in both groups, with no significant difference between groups (Figure 2b). There was also no significant difference in quality of life using the 5-Item EuroQol EQ-5D-5L Index Value Score^37,38^ (Figure 2c), the 100-point Visual-Analogue Scale (Figure 2d), or the WHODAS 2.0 Simple Score^39^ (Figure 2e). Similar patterns in these measurements were observed at secondary timepoints (30 days, 180 days, 360 days) (Supplemental Table 4).

**Table 3.**
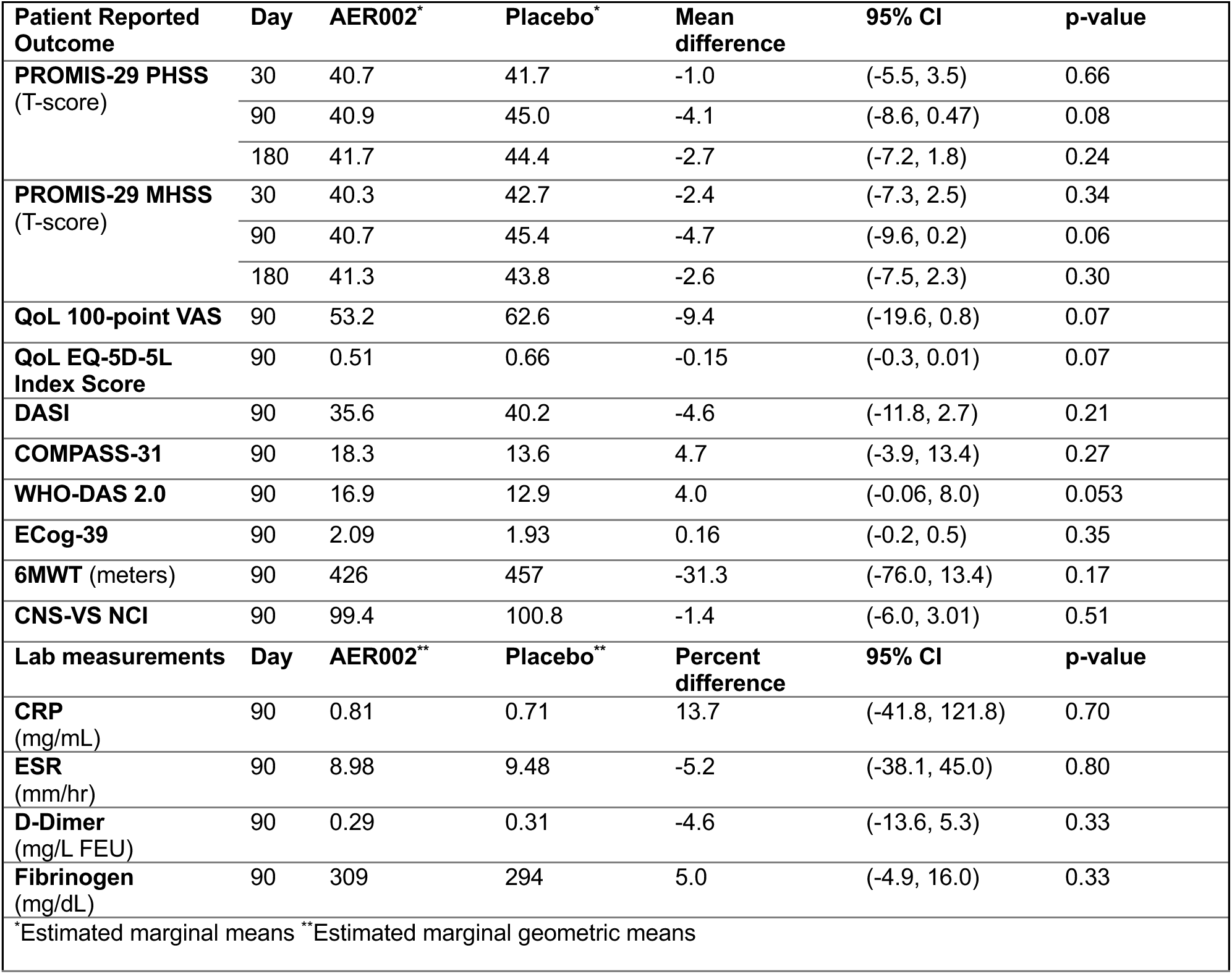
Linear mixed effect model results for continuous patient reported outcomes, performance measures and laboratory measurements.

**Figure 2.**
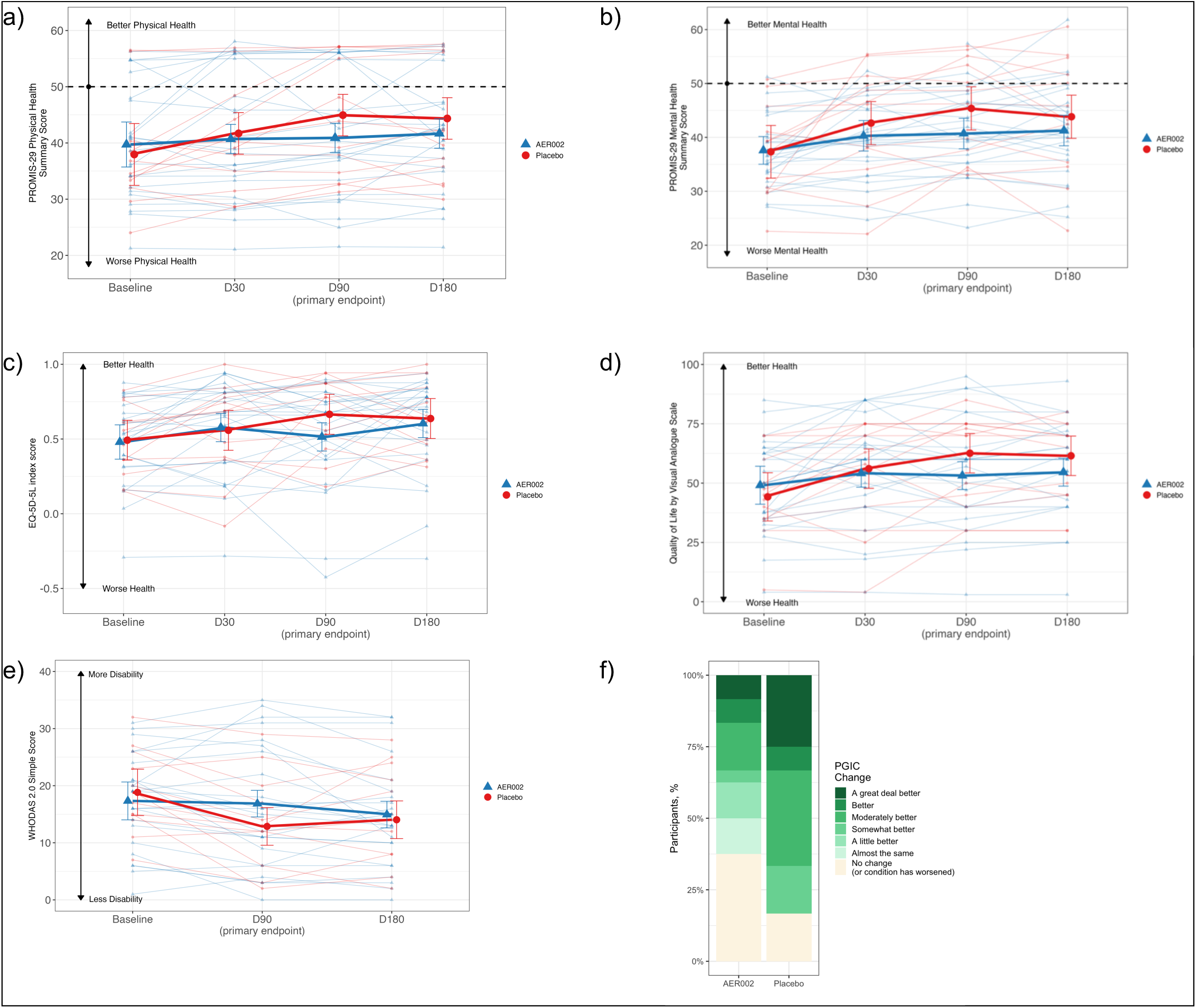
Changes in overall health. (a) PROMIS-29 Physical Health Summary Score. (b) PROMIS-29 Mental Health Summary Score. (c) EQ-5D-5L Quality of Life Index. (d) Quality of Life by Visual Analog Scale. (e) World Health Organization Disability Score. (f) Patient Global Impression of Change within treatment and placebo arms. Raw means are plotted for baseline and estimated marginal means are plotted for subsequent timepoints. Error bars indicate 95% CI.

#### Overall impression of change

Because of concerns that commonly used patient-reported outcome measurements may not adequately capture symptomatic changes in this patient population^40^, we used the Patient Global Impression of Change (PGIC) scale^41^ to directly query participants regarding whether they perceived any clinical changes following treatment and to identify self-reported responders and non-responders. Approximately half of the participants in the treatment arm and three-quarters in the placebo arm reported a perceived response to treatment (not significant; Figure 2f).

### Organ system-specific symptoms and performance measures

To address variability in Long COVID symptomatology between participants, we conducted in-depth assessments of several symptom domains among participants throughout the trial, utilizing a combination of validated patient-reported outcomes (PROs) and objective assessments.

#### Post-exertional symptoms

We did not observe a consistent pattern related to post-exertional symptoms exacerbated by the study; there was no difference in the odds of having consistent or resolving PEM between the treatment and placebo groups (Supplemental Figure 1). Among participants with PEM at baseline, five of 19 participants in the AER002 group and 4 of 10 participants in the placebo group reported improvement in PEM at Day 90 (p=0.45).

#### Cardiopulmonary symptoms

At baseline, the Duke Activity Status Index (DASI^42^) total score was similar between groups, reflecting impaired functional capacity (Supplemental Table 3). Similarly, distance achieved on a 6-minute walk test^43,44^ was comparable between groups. At Day 90, there were no significant differences in either DASI (Figure 3a) or 6-minute walk test (Figure 3b). Results were similar at secondary timepoints (30 days, 180 days, 360 days).

**Figure 3.**
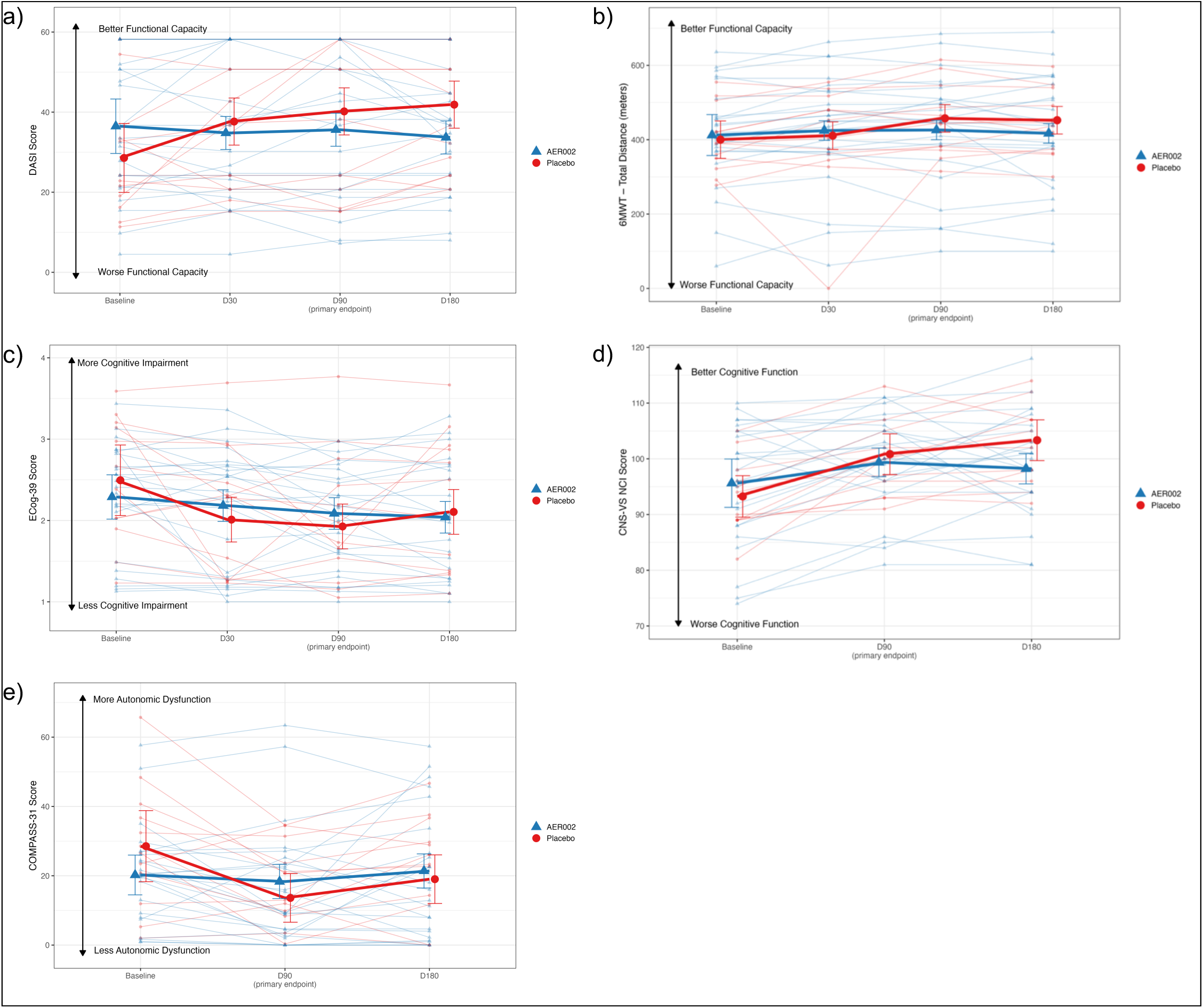
Changes in patient-reported and objective markers of physical activity, neurocognitive function, and autonomic function. (a) Duke Activity Status Index score. (b) 6-minute Walk Test total distance traveled. (c) Everyday Cognition-39 Score. (d) CNS-Vital Signs Neurocognitive Index. (e) Composite Autonomic Symptom Score. Raw means are plotted for baseline and estimated marginal means are plotted for subsequent timepoints. Error bars indicate 95% CI.

#### Neurocognitive symptoms and performance

At baseline, self-reported (Everyday Cognition Scale (ECog^45^)) and objective (CNS-Vital Signs^46^) neurocognitive performance was similar between groups (Supplemental Table 3). At Day 90 following the infusion, there were no significant differences in either ECog (Figure 3c) or CNS-Vital Signs neurocognitive index score (Figure 3d). Results were similar at secondary timepoints (30 days, 180 days, 360 days).

#### Autonomic symptoms and performance

At baseline, self-reported autonomic dysfunction via the Composite Autonomic Symptom Score (COMPASS-31^47^) instrument was similar between groups (Supplemental Table 3); on active stand test^48^, one patient met the criteria for objective autonomic dysfunction. At Day 90 following the infusion, there were no significant differences in either COMPASS-31 (Figure 3e) or active standing test (one positive). Results were consistent at secondary timepoints (30 days, 180 days, 360 days).

### Laboratory measurements

#### Targeted blood-based measures of inflammation, immune activation, and clotting

We did not identify statistically significant or clinically meaningful differences in targeted measurements of inflammation (C-reactive protein, erythrocyte sedimentation rate), immune activation (IL-6, TNF-alpha, interferon-gamma, IL-1beta), or clotting (D-dimer, fibrinogen) between groups at baseline (Supplemental Table 3) or any subsequent timepoint (Figure 4a-h).

**Figure 4.**
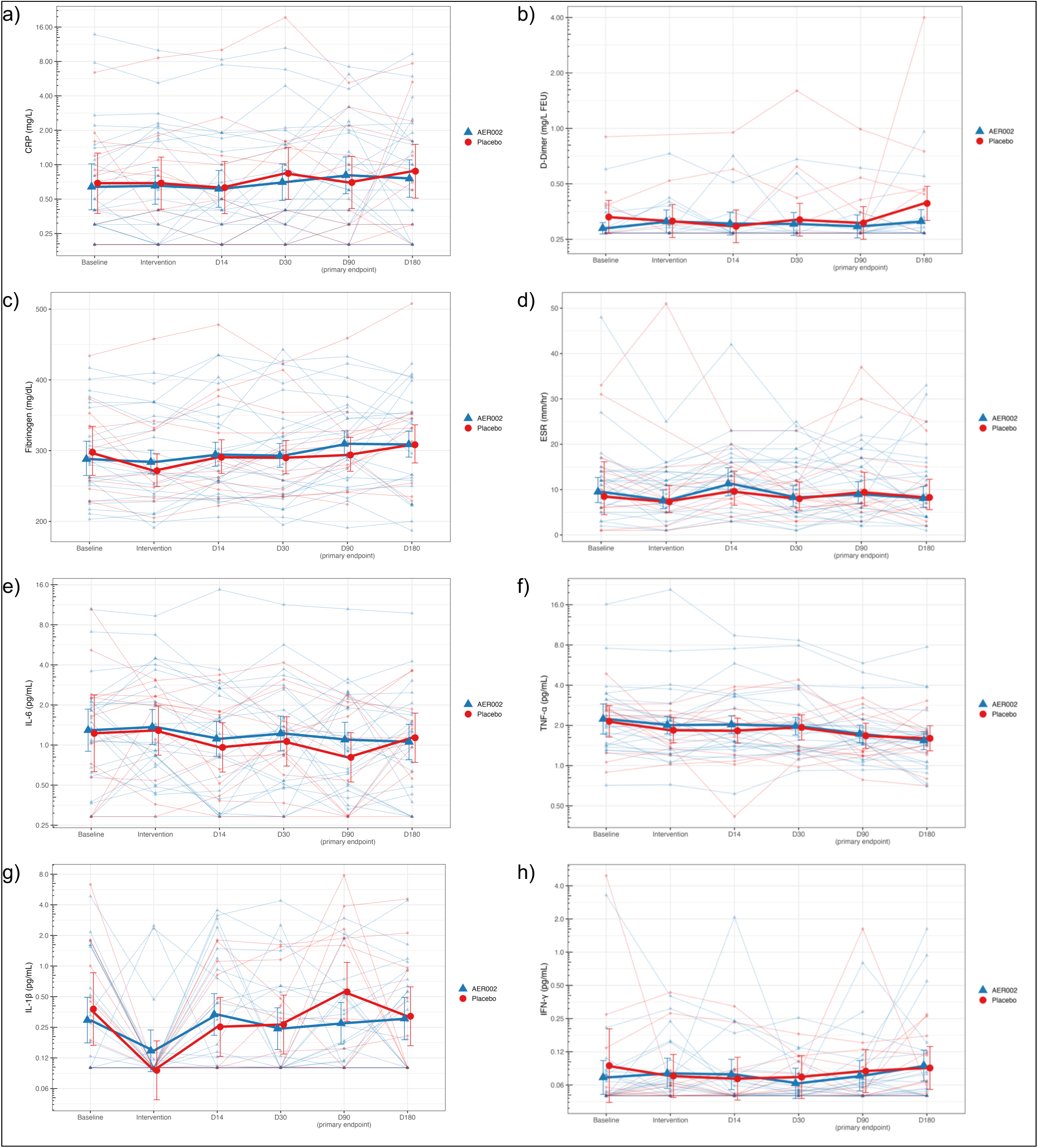
Changes in laboratory-based biomarkers. (a) C-reactive protein. (b) D-Dimer. (c) Fibrinogen. (d) Erythrocyte sedimentation rate. (e) Interleukin-6. (f) Tumor necrosis factor-alpha. (g) Interleukin 1-beta. (h) Interferon-gamma. Raw means are plotted for baseline and estimated marginal means are plotted for subsequent timepoints. Error bars indicate 95% CI.

#### Antibody measurements and AER002 pharmacokinetics (PK)

Baseline anti-SARS-CoV-2 antibodies were measured by single-molecule array^49^. Anti-spike (anti-S), anti-spike S1 (anti-S1), and anti-receptor binding domain (anti-RBD) antibody levels tended to be higher in placebo versus AER002 arms at baseline (anti-S [median (IQR): placebo 124 (84.1 – 165) vs AER002 62.3 (51.2 – 111)] p=0.026, anti-S1 [median (IQR): 224 (144 – 311) vs 136 (56.4 – 191)] p=0.032, and anti-RBD [median (IQR): 170 (43.1 – 233) vs 82.6 (30.8 – 126)] p=0.072), while anti-nucleocapsid (anti-N, [median (IQR): 3.50 (1.96 – 7.33) vs 2.28 (1.79 – 4.01)]) antibody levels were not significantly different (p=0.54) (Figure 5).

**Figure 5.**
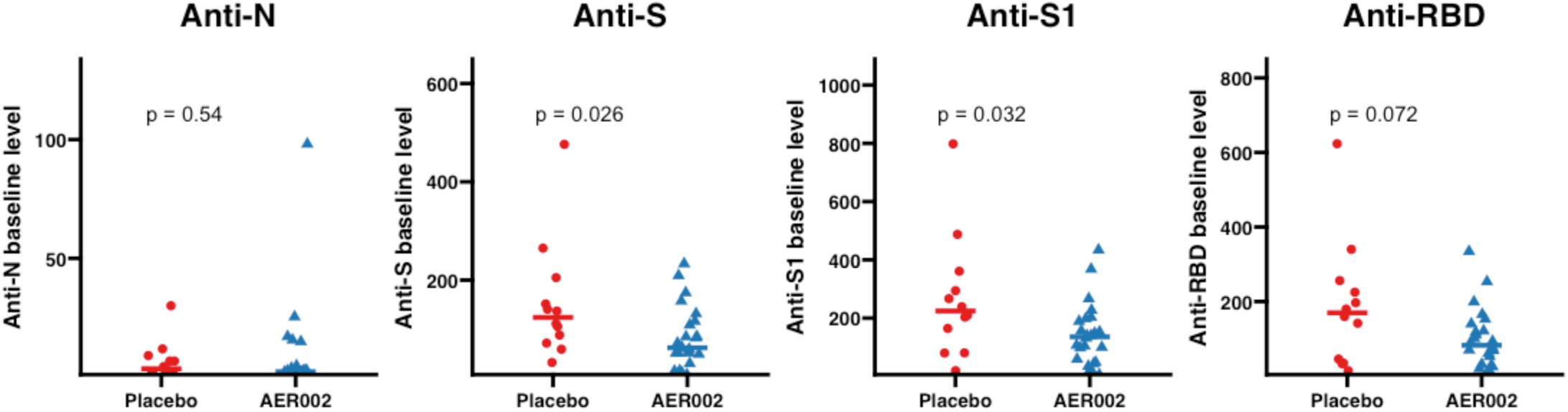
Baseline levels of SARS-CoV-2 antibodies. (a) Anti-nucleocapsid. (b) Anti full-length spike. (c) Anti-S1. (d) Anti-receptor binding domain.

AER002 serum concentration was measured via a validated LC-MS/MS method quantifying a unique CDR1-region signature peptide, based on a previously described method^31^. AER002 PK was characterized using noncompartmental and population pharmacokinetic approaches. A two-compartment population pharmacokinetic model (Supplemental Table 5) with between-participant variability for clearance and central volume of distribution was constructed using 114 total serum concentrations from 24 participants obtained over 6 months. Although not statistically significant, AER002 levels tended to wane more rapidly in female participants (terminal half-life 75.5 [median (IQR): 85.4 (65.6-110.3)] vs. 107.8 [median (IQR): 107.1 (81.1-140.1)] days for female vs. male, respectively, p=0.28). In population PK analyses, male participants were estimated to have 1.96-fold (95% CI: 1.27-2.88) higher peripheral distribution volume than female participants.

#### Plasma proteomics and pathway analysis

Plasma proteomic profiling was performed using the Olink® Reveal platform (Olink® Proteomics, Uppsala, Sweden), a highly sensitive and specific proximity extension assay (PEA)–based technology. A total of 1,034 unique proteins were quantified from cryopreserved plasma samples collected at baseline and on the day of infusion prior to administration of study drug (averaged to represent baseline values), and at 14 and 90 days post-infusion^50^. We calculated unadjusted p-values and beta estimates and considered proteins with p<0.05 and beta estimate +/- 0.3, consistent with an approximate 1.25-fold change, to be potentially clinically meaningful. We also indicated proteins that met the threshold for statistical significance on multiple testing using the Storey Q method^51^.

Baseline proteomic profiles were largely comparable between treatment groups, with only minor differences observed for a small number of proteins prior to intervention (Supplemental Figure 2). At Day 14 (Figure 6a), treatment was associated with modest proteomic differences relative placebo, affecting a small subset of markers linked to immune regulatory signaling and tissue remodeling processes (BTLA, SPIK6, MYL3). At Day 90 (Figure 6b), we observed a broader pattern of proteomic differences; a larger number of markers showed differences in unadjusted analyses, with the majority demonstrating negative beta estimates indicative of lower levels in the AER002 group compared to placebo. These proteins included multiple markers involved in immune activation, inflammatory signaling, and stress responses pathways, including proteins associated with interferon responses and innate immune responses (GBP2, IGBP1), apoptosis and regulation of cell death (CASP7, BAX, CSDMD, DFFA), and intracellular signaling pathways such as MAP kinase and NF-kappa B signaling (MAP2K1, NFKBIE, IRAK4, AKT1S1). Only a small number of proteins demonstrated increased levels with AER002 (PTPN14, a regulator of cell signaling). However, Molecular Signatures Database (MSigDB) Hallmark gene set collection analyses^52^ did not reveal obvious immune or inflammatory pathway trends between groups, likely due to limited sample size.

**Figure 6.**
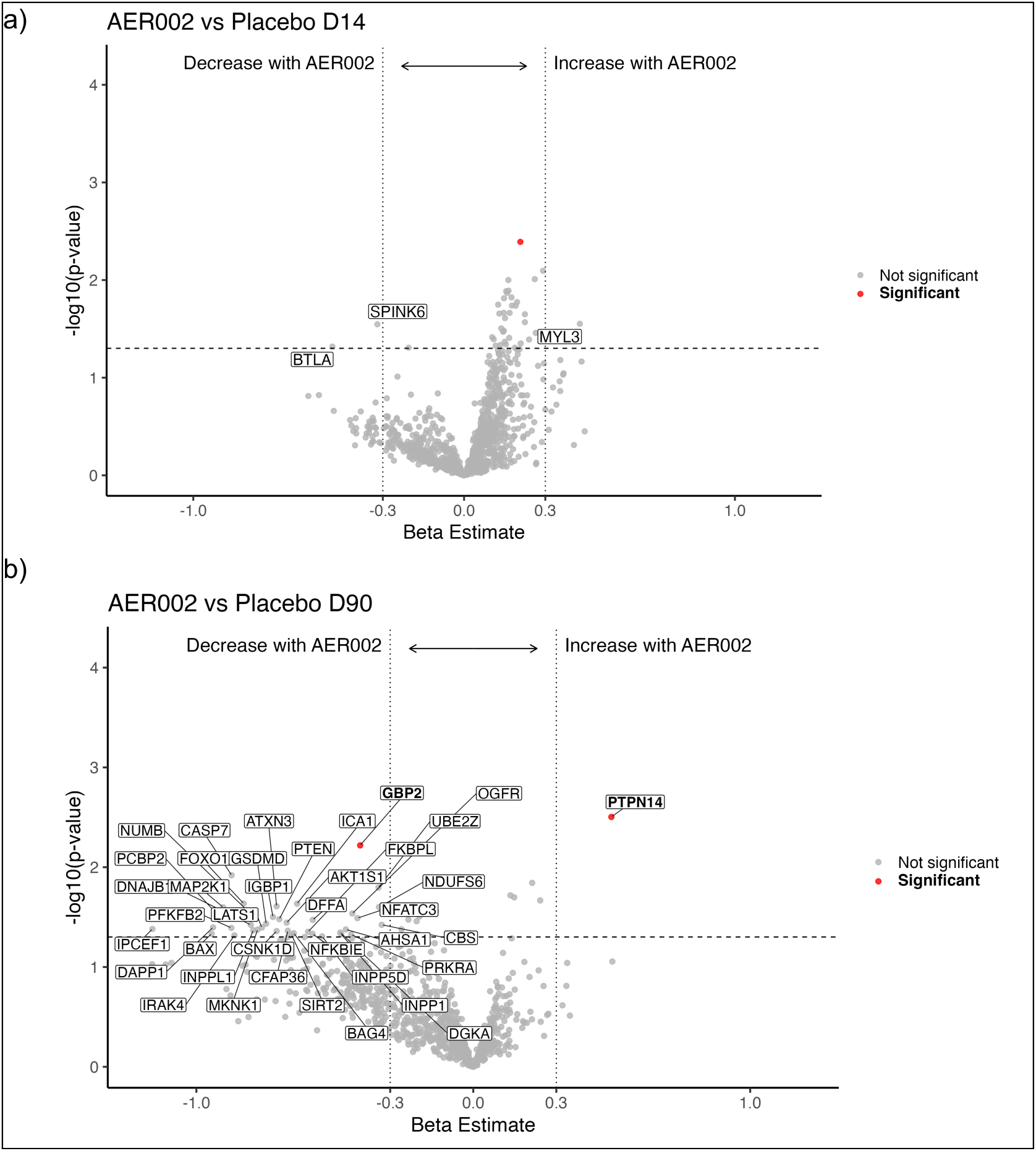
Plasma proteomics. (a) AER002 vs Placebo at D14. (b) AER002 vs Placebo at D90. Significant proteins with q-value < 0.05 are shown in bold red. The horizontal dashed line indicates the unadjusted p-value significance threshold (p = 0.05). Vertical dashed lines indicate beta estimates of -0.3 or 0.3.

### Per protocol analysis

We conducted a per-protocol analysis in which we excluded individuals with confirmed and suspected on-study reinfections before the primary endpoint. The results were largely unchanged. There were no significant p-values for all measures reported in detail above including physical health and quality of life outcomes, overall impressions of change, and organ-system specific symptoms and performance measures (Supplemental Table 6).

### Patient ascertainment of treatment

As part of the observational cohort study in which all participants were co-enrolled, we assessed patient ascertainment of treatment group assignment shortly before unblinding. Briefly, participants were asked to predict the group to which they had been assigned (AER002 vs placebo) and rate their confidence in their answer on a scale from 1 (no confidence) to 5 (complete confidence). In the AER002 group, 42% predicted their treatment assignment correctly, compared to 50% in the placebo group. For both groups, most rated their confidence as moderate to low (<=3) on the confidence scale.

### Optional procedures

Concurrent with trial activities, all participants were offered the opportunity to complete optional procedures including flexible sigmoidoscopy with colorectal sampling, positron emission tomography-computed tomography (PET-CT) imaging, and cardiopulmonary exercise testing (CPET). Participants were consented separately to these procedures, which were completed under independent protocols at baseline and approximately three months post-infusion. To avoid biasing the main study results, in all cases, the post-infusion optional procedures occurred on a separate day following the assessment of primary and secondary outcomes at the 90-day timepoint.

#### Tissue viral persistence and inflammation

We and others previously demonstrated that gastrointestinal tissue can harbor persistent SARS-CoV-2 single- and double-stranded RNA^6,15,16^ and, given the immune regulatory nature of this environment, may support ongoing viral activity as a driver of systemic inflammation. In a subset of participants (n=17; 12 AER002, 5 placebo), we performed flexible sigmoidoscopy with colorectal sampling a median of 10 days (range 1-29) prior to infusion and again a median of 98 days (range 85-195) following infusion. An additional participant (who received AER002) completed the initial collection but declined repeat sample collection.

We used multiplexed chromogenic RNAscope to identify SARS-CoV-2 single-stranded spike RNA and double-stranded orf1ab RNA in formalin-fixed paraffin embedded tissues, with reference to control tissue collected prior to the pandemic (Figure 7a). Prior to infusion, we detected single-stranded spike RNA in one participant (718 days from most recent known infection; Figure 7b). This participant received AER002 and reported significant improvement over the course of the study; no RNA was detected on their post-infusion biopsy. Following infusion, we detected single- and double-stranded spike and ORF1ab RNA in three participants (all received AER002) who did not have detectable RNA at baseline sampling (Figure 7c). Of note, all three had known reinfections following the infusion (biopsies were a median of 21 days from most recent infection); two additional participants (both received placebo) with intervening reinfections did not have signal detected on follow-up biopsies 18 and 57 days after reinfection (Supplemental Table 7). No definitive SARS-CoV-2 RNA was observed in RNA from bulk colorectal tissue by qPCR (spike, nucleocapsid).

**Figure 7.**
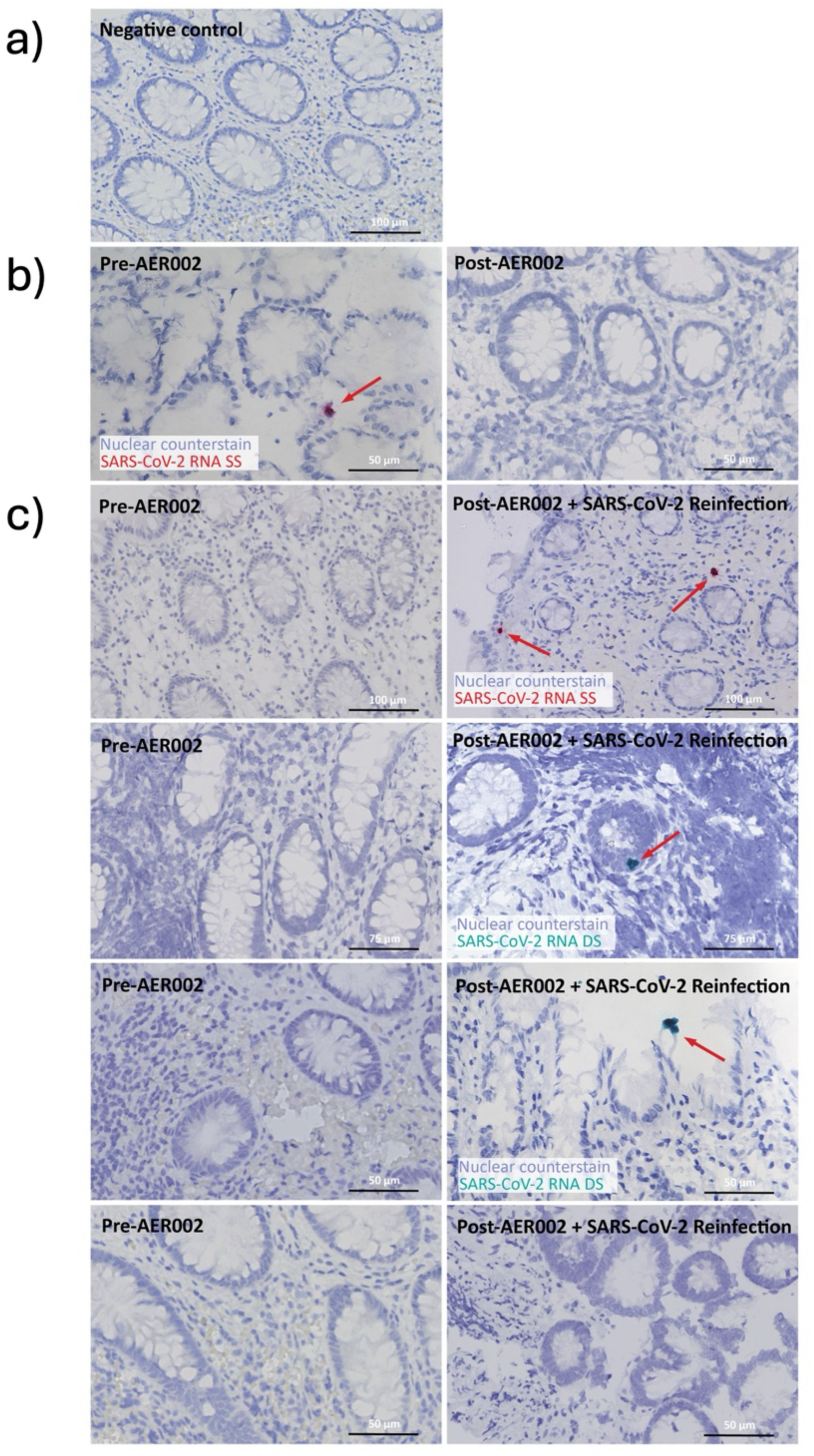
Results from optional gut biopsies. (a) Control (pre-pandemic) gut tissue. (b) Detection of SARS-CoV-2 RNA in one participant at baseline, without detectable RNA at follow-up. (c) Detection of SARS-CoV-2 RNA at follow-up in 3 of 5 participants with known re-infections following the infusion. No detection of SARS-CoV-2 RNA at follow-up in 1 of 5 participants with known re-infections following the infusion.

In addition, we used the custom nCounter human gene and SARS-CoV-2 RNA quantitation assay to simultaneously measure over 750 human transcripts involving genes associated with viral immune responses and nine transcripts across the SARS-CoV-2 genome in colorectal tissue samples. Though sample sizes were limited due to exclusion of participants with interim SARS-CoV-2 reinfection and those with poor specimen quality, there were no apparent differences in genes associated with viral infection and inflammation between drug (n=9) and placebo (n=2) arms following treatment. Using this method, we also did not observe significant differential gene expression between baseline and post-infusion timepoints in either the placebo or AER002 arm.

#### PET imaging analysis

We previously identified differences in whole-body T cell activation states in people with and without prior COVID-19 by non-invasive PET-CT imaging using a novel radiotracer ([18F]F-AraG)^15^. [18F]F-AraG is phosphorylated and selectively retained in the mitochondria of activated T cells and is therefore highly specific for cycling CD8+ T cells and, to a slightly lesser extent, CD4+ T cells. While myeloid immune cell uptake is also possible, it is far lower than what is observed in T cells. Here, we performed [18F]F-AraG PET imaging in a subset of participants a median of 10 days (range 4-151) prior and 98 days (range 83-179) following study drug (AER002; n=6) or placebo (n=4) and compared mean standardized uptake values (SUVmean) across tissue regions of interest (ROI) that we identified in our prior observational study.

In cross-sectional analyses prior to infusion, we observed significantly lower cauda equina (lumbar spinal cord) uptake in those who received AER002 compared to placebo but no other significant differences across other ROIs (Figure 8a). Following treatment, however, we detected significantly lower tracer uptake in those who received AER002 compared to placebo in several tissues, including temporal brain lobes, thoracic spinal cord, cauda equina, and submandibular glands (all p<0.05; (Figure 8a). In the context of sample size limitations and inter-participant variations in tracer uptake, we did not observe any significant longitudinal differences between baseline and post-infusion images within AER002 and treatment groups but did observe a trend toward decreased uptake in submandibular glands in the treatment arm in contrast to an increase in uptake in the placebo arm (Figure 8b-c).

**Figure 8.**
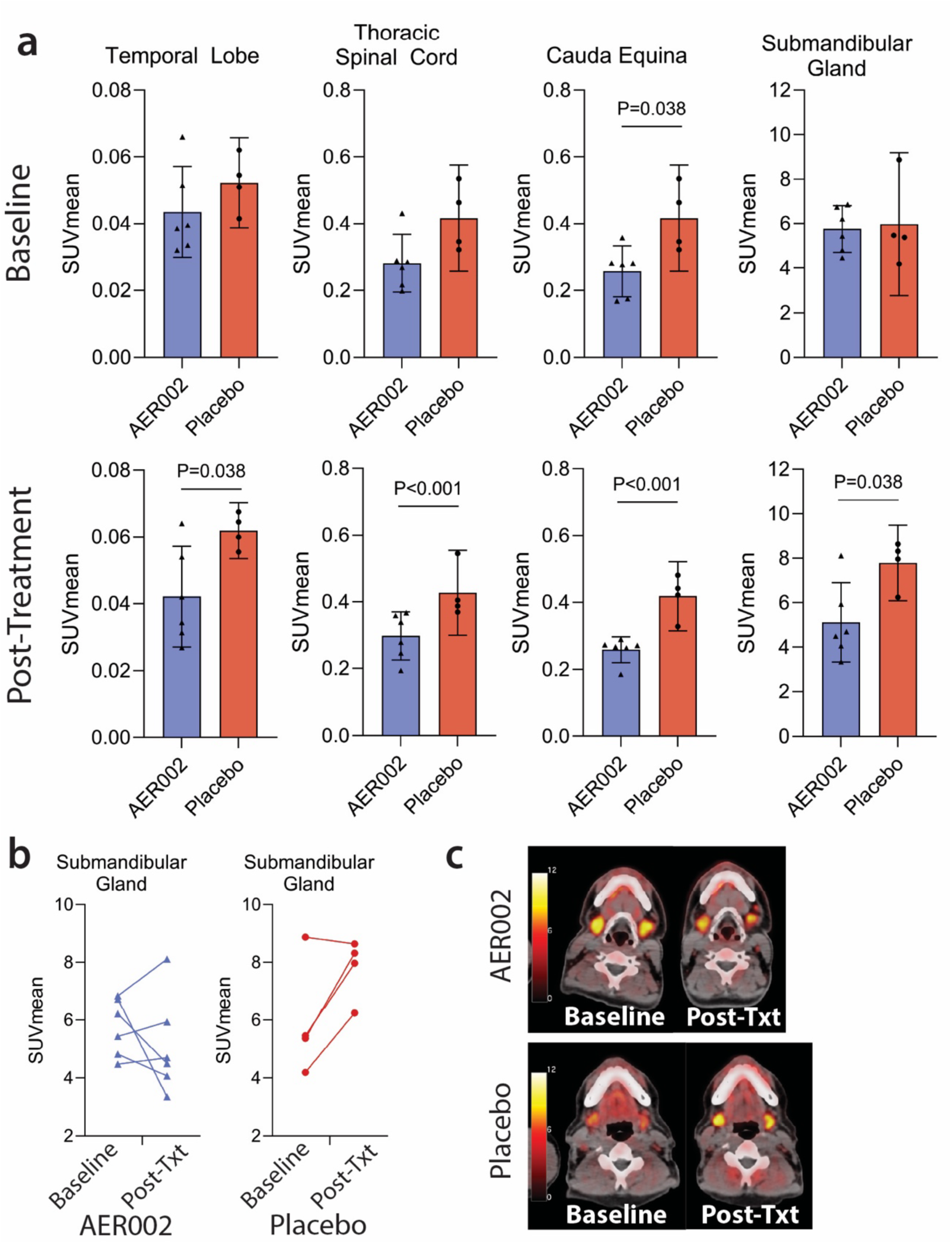
Results from optional [18F]F-AraG PET-CT imaging. (a) Baseline and post-infusion mean standardized uptake values (SUV) in selected regions of interest (ROI). (b) Within-participant changes in SUV in the submandibular glands baseline and post-infusion. (c) Representative imaging of the submandibular glands from participants in each arm

#### Cardiopulmonary performance

A subset of participants completed opt-in cardiopulmonary exercise testing (CPET) at baseline (n=9) and following the 3-month timepoint (n=5). At baseline, the mean peak oxygen consumption was 25.3 ml/kg/min, or 88% predicted by the Wasserman equations, at a mean power output of 149 Watts. Among those who received AER002 (n=4), all tests were to maximal effort (respiratory exchange ratio >1.16). There was no change in peak oxygen consumption from baseline to 3-months (−0.58 ml/kg/min difference (slightly worse) p=0.60), peak oxygen consumption on the percent predicted scale (0.5% increase, p=0.92), or peak work (2.25 Watts increase, p=0.53).

### Exploratory analysis of self-reported “responders”

To better understand the potential biological effects of AER002 in people with Long COVID, we conducted pre-specified exploratory analyses of self-identified responders in both treatment arms. We defined “responders” as those who reported an improvement on the PGIC scale of “Moderately Better” or more at the 90-day and/or 180-day timepoints. Twelve participants (6/24 AER002, 6/12 placebo) met criteria at both 90 and 180 days. Four participants (2/24 AER002, 2/12 placebo) met criteria at 90 days only while three participants (2/24 AER002, 1/12 placebo) met criteria at 180 days only (Figure 2F).

#### Impact of inflammation, immune activation, and clotting markers on self-reported treatment response

Baseline fibrinogen levels were higher in responders compared to non-responders in the AER002 group (324 vs 258 mg/dL, p=0.048). A similar trend was observed in the placebo group but did not meet statistical significance. We did not identify statistically significant differences in baseline CRP, ESR, or baseline cytokines (i.e., IL-6, TNF-alpha, interferon-gamma, IL-1 beta) between responders and non-responders.

#### Proteomic signatures of self-reported treatment response

Responders to AER002 at Day 90 exhibited modest but selective proteomic differences relative to non-responders, characterized primarily by lower levels of several signaling and inflammatory-associated proteins (Supplemental Figure 3). Proteomic differences between responders and non-responders were also observed in the placebo arm (Supplemental Figure 4), indicating that response-associated protein changes were not unique to AER002 treatment, although the individual proteins and pathways were different.

#### Impact of SARS-CoV-2 antibody levels on self-reported treatment response

Among the entire study population, responders had significantly higher baseline levels of anti-S (median normalized values for responder vs non-responder: 127 vs 60.5, p=0.033), anti-S1 (207 vs 106, p=0.036), and anti-RBD (151 vs 65.1, p=0.033) antibodies, but not baseline anti-N (2.93 vs 2.05, p=0.178) (Figure 9a). In both the AER002 and placebo groups, having high levels (classified by baseline levels above median value) of anti-S, anti-S1, anti-RBD, and anti-N antibodies was associated with greater odds of being a responder (in each of 4 different logistic regression models for each antibody type). Due to low sample size and high correlation between antibody types we were unable to include more than one antibody in a single model (Supplemental Table 5).

**Figure 9.**
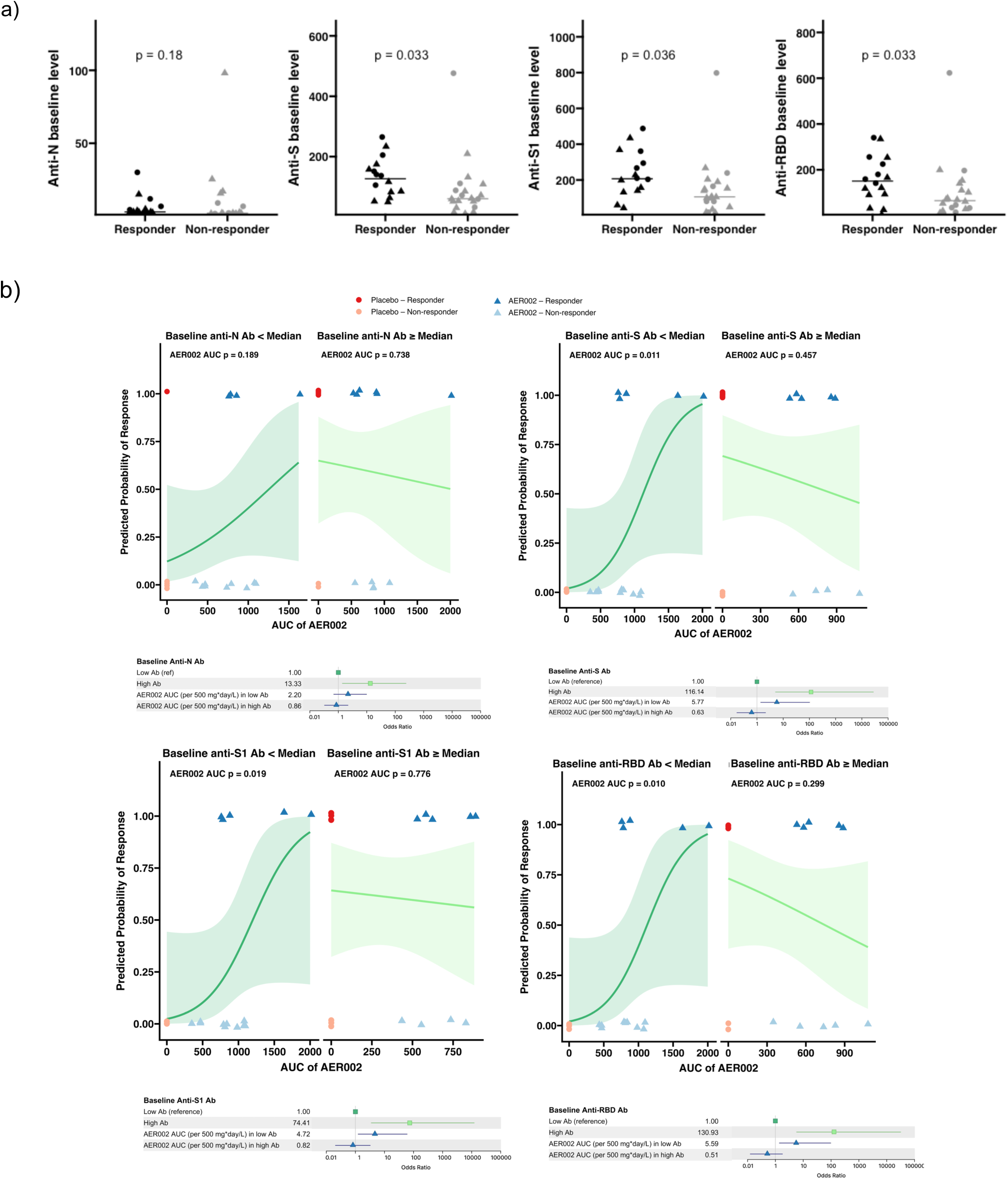
Impact of baseline anti-SAR-CoV-2 and AER002 pharmacokinetics on self-reported response. (a) Comparison of baseline anti-SARS-CoV-2 antibodies between self-reported responders and non-responders. (b) Multiple logistic regression of each baseline antibody level and AER002 exposure (AUC: area under the concentration-time curve) on likelihood of self-reported response displayed in pairs of logistic regression plots (Left: baseline antibody < median, Right: baseline antibody ≥ median) for each antibody (clockwise from top left: anti-N, anti-S, anti-RBD, anti-S1). Forest plots of odds ratios (OR) derived from each regression analysis depicted below each pair of plots.

We also assessed an interaction between AER002 AUC and levels of each of the antibody types in these regression models. A greater AER002 AUC was associated with greater odds of being a responder in participants who exhibited low baseline anti-S, anti-S1, and anti-RBD levels (less than median cutoffs) (Figure 9b). Specifically, for each 100 mg*day/L AER002 AUC increase, there was 1.41-fold higher odds of response (95% CI, 1.07-2.50, p=0.011) for participants with low baseline anti-S, 1.36-fold higher odds (1.04-2.27, p=0.019) for anti-S1, and 1.41-fold higher odds (1.07-2.50, p=0.010) for anti-RBD. For those with low anti-N, increasing AER exposure was not associated with greater odds of response (OR 1.17 [0.93-1.58], p=0.189). An association between odds of being a responder and AER002 AUC was not found in the subgroup who had high baseline antibodies.

## DISCUSSION

Although numerous studies now suggest that SARS-CoV-2 can persist in some people following COVID-19^6^, the precise form, location, and mechanism of persistence remain incompletely understood^17^. Several trials of small molecule antivirals such as nirmatrelvir/ritonavir administered for 15 days have failed to show a benefit in Long COVID^21,22^, perhaps due to short duration of treatment or the possibility that SARS-CoV-2 persistence may not represent active viral replication^5,17^. We hypothesized that treatment with a long-acting SARS-CoV-2-specific monoclonal antibody would address these potential issues and result in symptomatic improvement either through direct neutralization of spike protein or the clearance of infected cells through processes mediated by retained effector function. However, across a wide range of patient-reported and objective outcomes, we found no group-level benefit of monoclonal antibodies compared with placebo in people with clinically defined Long COVID. Despite the negative result, mechanistic subgroup analyses provide insight into the biology of Long COVID and can inform the design of future trials.

SARS-CoV-2 persistence has been documented in the post-acute phase of COVID-19 but is not specific to Long COVID. Persistent SARS-CoV-2 may also not be a driver of disease for all patients with the condition; rather, a subset of patients with Long COVID may harbor persistent virus. Estimates of SARS-CoV-2 persistence vary from 10-40% in the months following COVID-19; less is known about persistence on the order of years^6,17^. The lack of a validated, accessible biomarker to confirm the presence of SARS-CoV-2 persistence presents a major challenge to the conduct of clinical trials targeting this mechanism^5^. At the time of our study, no such measure was available. Thus, we did not recruit participants based on a metric of SARS-CoV-2 persistence, which could be one reason the study did not meet the primary outcome. Tissue measurements have been proposed as a better metric for assessing viral persistence^17^, but only one of the participants who underwent gut biopsy had detection of SARS-CoV-2 RNA in gut tissue at baseline; notably this individual reported improvement. As a result, it is possible that this small study simply did not assess enough individuals with the potential to benefit from this therapeutic approach. Coordinated efforts to develop and validate biomarkers of viral persistence in more accessible matrices like blood are needed; once available, future studies should consider using such measurements to determine eligibility.

Participants in this trial had been experiencing Long COVID for about two years, considerably longer than the period in which SARS-CoV-2 persistence has thus far been linked to Long COVID symptoms (3-14 months)^7–9,53^. It is possible that SARS-CoV-2 persistence drives disease early on, but gives way to mechanisms like chronic immune dysfunction, autoimmunity, vascular injury, or mitochondrial dysfunction in a later post-acute phase. Longitudinal studies have suggested that the biology of ME/CFS, another multi-system disorder often attributed to an infectious trigger, could evolve over time^54,55^. Larger studies should consider analyses stratified by time since the Long COVID-triggering SARS-CoV-2 infection to assess whether interventions may have differential benefit in various stages of the disease process.

We made several observations in our exploratory analyses that may inform future work. Participants in both groups who reported improvement tended to have higher anti-SARS-CoV-2 antibody levels at baseline, which suggests that such individuals have the potential to exhibit spontaneous improvement; consistent with this observation, our pharmacokinetic analysis revealed that increasing exposure of AER002 did not increase odds of response among those with high baseline antibody levels. In contrast, participants with low baseline anti-SARS-CoV-2 antibody levels had lower odds of spontaneous response, and higher exposures of AER002 were associated with increased odds of improvement among this group. Together, these observations suggest that antibody therapy or other immune enhancing approaches might work best in those who exhibit a weak immune response to the virus, and that higher doses or more sustained administration of anti-SARS-CoV-2 antibody interventions may increase the likelihood of response. These findings warrant future studies that screen participants based on baseline antibody levels and/or dose modern monoclonals using strategies that optimize anti-SARS-CoV-2 antibody exposure (e.g., repeat dosing).

Long COVID is associated with systemic inflammation^56^, and our prior PET imaging work has revealed tissue-based immune activation in multiple organ systems^15^. Although underpowered, exploratory PET imaging of immune cell activation demonstrated significantly lower SUV in brain, thoracic spinal cord and, most notably, the submandibular gland in participants following AER002 treatment in comparison to placebo. Tracer uptake (denoting T cell activation) in the submandibular glands decreased in most participants in the AER002 group and increased in the placebo group, which may suggest monoclonal antibody-mediated blunting of immune activation potentially driven by an effect on SARS-CoV-2 reservoirs in oral glandular tissues. In conjunction, our exploratory proteomics analyses suggested that AER002 treatment could be associated with a pattern consistent with attenuation of multiple immune and cellular activation pathways, consistent with evolving pharmacodynamic effects following treatment administration. While these data are not definitive proof of AER002 modifying immune activation states via antiviral activity, they suggest that the AER002 had a biological effect and can potentially alter pathways related to inflammation and immune function that could be related to Long COVID.

As in other Long COVID trials ^21,22,57–59^, we observed a robust response among those receiving placebo. Such findings warrant discussion, as they may inform future trial design. The placebo group exhibited higher baseline SARS-CoV-2 antibody levels than the AER002 group, which we found to be a strong predictor of improvement; this may have driven improvement in the placebo arm and blunted a therapeutic effect in the treatment arm. It is also possible that improvements in both arms were driven by changes due to intensive observation (i.e., a Hawthorne effect^60^), the belief that the specific intervention being studied would be beneficial (i.e., a placebo effect^61^), or idiosyncratic improvement related to the use of concomitant medications (although participants were asked to minimize medication changes during the study). If a placebo effect occurred, it should be noted that these responses are biologically meaningful, engaging neuroimmune networks that influence both symptom perception and inflammatory processes^62^. Such responses can alter pain signaling, autonomic function, and inflammation^63,64^. Studies in inflammatory arthritis have demonstrated that learned or expectancy-driven mechanisms can modulate disease activity and even objective inflammatory markers^65^. In this light, placebo responses in Long COVID trials may reflect activation of similar regulatory circuits linking the brain and the immune system. Regardless of the cause of the improvement observed in the placebo arm, it is notable that few participants reported that they had returned to their pre-COVID baseline. From a clinical standpoint, these effects highlight the profound interconnectedness of mind, brain, and body in chronic illness^66^. From a design standpoint, strategies such as standardized treatment messaging, placebo run-ins, or crossover designs may help distinguish true therapeutic effects from expectancy-related improvements and temporal effects^67^; much larger studies that are powered around this potential for improvement are also needed.

Strengths of this study include robust phenotyping of participants with both patient-reported and objective measures, its placebo-controlled design, long duration of follow-up, inclusion of detailed pharmacokinetic analyses, and the inclusion of optional procedures to support pathogenesis research. There are several notable limitations. The sample size was small and the 2:1 randomization resulted in a small placebo arm. We recruited participants who were able to attend frequent, prolonged visits at the medical center and thus excluded those with the most severe illness (e.g., those who were homebound). As noted above, we could not conduct biomarker screening to determine eligibility. Although participants attributed their symptoms to a preceding test-confirmed SARS-CoV-2 infection and clinician assessment ruled out other clear etiologies, it remains possible that a portion of the study population had symptoms caused by conditions other than Long COVID. Although we recruited participants with Long COVID attributed to variants against which AER002 had activity, one third of the cohort had experienced a SARS-CoV-2 reinfection after the date at which AER002 was expected to have lost neutralizing capacity; the potential for so-called “mixed reservoirs” has been suggested^17^. Although participants were requested to not make alterations to their background medication regimens during the study, there was significant use of concomitant medications that could have affected certain outcome measures and led to general trends towards improvement across both study arms. Additionally, many of our outcome measures have not been specifically validated in Long COVID and may not have adequately captured all aspects of the disease^55^.

Finally, although our hypothesis is that SARS-CoV-2 persistence is the principal driver of Long COVID, it is possible that other pathways implicated in the disease – such as immune dysfunction – may actually be a cause of SARS-CoV-2 persistence rather than a consequence of persistence. In such a case, immune modulating interventions rather than those targeting the virus may be warranted^5^. Such studies are now underway.

In summary, we did not identify a group level benefit of AER002 in comparison to placebo for the treatment of Long COVID. Despite the negative result, we made several observations that suggest promise for monoclonal antibody therapies in Long COVID and should inform the design of future studies. Future trials targeting viral persistence in this heterogeneous condition would benefit from confirmation of SARS-CoV-2 persistence at the time of enrollment, larger sample sizes to mitigate heterogeneity and reinfection, utilization of agents with broad activity against both historical and modern variants, and consideration of screening of participants for immune phenotypes (e.g., low baseline antibody levels) more likely to benefit from treatment and/or dosing strategies that achieve sustained therapeutic levels for a significant duration of time.

## METHODS

### Participants

Participants were recruited from the University of California, San Francisco (UCSF)-based Long-term Impact of Infection with Novel Coronavirus (LIINC) cohort (NCT04362150)^68^, which enrolls and prospectively measures adults in the post-acute phase of SARS-CoV-2 infection. All participants were required to enroll in LIINC prior to screening for this trial. Briefly, interested LIINC participants completed a screening visit at which their eligibility for this trial was assessed. The trial enrolled adults (≥18 years) with clinical evidence of Long COVID, defined as at least two moderate or severe symptoms that were new or worsened since the time of SARS-CoV-2 infection and were not known to be attributable to another cause, which had been present for at least 60 days following a test-confirmed SARS-CoV-2 infection that occurred in the period in which AER002 was expected to have neutralizing activity against circulating variants (i.e., prior to August 15, 2022). Our definition was based on the World Health Organization consensus definition^69^; although not available when the study was planned, our definition is consistent with that which has since been put forth by the National Academies of Sciences, Engineering, and Medicine^1^. We excluded those who had ever previously received a SARS-CoV-2-specific monoclonal antibody, had received COVID-19 convalescent plasma or a SARS-CoV-2 vaccine within the preceding 60 days, who had major surgery within the prior 6 months, had medical comorbidities including active cardiovascular disease (defined as recent myocardial infarction or stroke, or history of congestive heart failure or pulmonary arterial hypertension), viral hepatitis, uncontrolled HIV infection, coagulopathy, severe anemia, moderate or severe immunocompromise according to the 2023 CDC COVID-19 guidelines^70^, and those with a history of anaphylaxis or similar significant allergic reaction. We also excluded those with a diagnosis of ME/CFS or dysautonomia that preceded their SARS-CoV-2 infection. The full list of eligibility criteria is in Supplementary Table 8.

### Trial Design

Following confirmation of eligibility, participants underwent a series of baseline assessments. On the day of the infusion, they were randomized in a 2:1 ratio to receive a single intravenous infusion of study drug or placebo, which was prepared by an unblinded pharmacist. The participant and study team were blinded to treatment allocation. Participants underwent follow-up evaluations at approximately Day (D) 14, D30, D90, D180, and D360 following the infusion. The study team and participants remained blinded until the database lock following the D180 visit for all participants, after which participants were followed until D360 in an open-label manner to assess long-term safety outcomes.

The primary objective of the study was to evaluate the safety and tolerability of AER002 in individuals with Long COVID. The primary safety endpoint was the number of participants in each arm who experienced a grade 3 or greater adverse event that was determined to be definitely, probably, or possibly related to study treatment. The primary efficacy endpoint was the PROMIS-29 Physical Health Summary Score between Baseline and D90. There were multiple pre-specified secondary endpoints as outlined in Supplementary Table 9. Exploratory measures included pharmacokinetic measures and analyses of various biomarkers.

### Study Drug

Participants received a single intravenous infusion of AER002 1200mg or matching placebo. The placebo was comprised of the formulation buffer for AER002, which included 20mM histidine, 8% sucrose, and 0.04% Polysorbate 80. For this study, the assigned product was admixed in 250mL of normal saline and infused over 30 minutes.

### Clinical Measurements

#### Patient Reported Outcomes

##### PROMIS-29

Participants completed the self-administered Patient-Reported Outcomes Measurement Information System (PROMIS)-29 v2.1 survey to assess broad functional ability and quality of life^34^. The PROMIS questionnaires are scaled to fit a T-scale, with a normal distribution including a mean score of 50 and a standard deviation of 10 points. PROMIS-29 scales spanning seven health domains (physical function, fatigue, pain interference, depressive symptoms, anxiety, ability to participate in social roles and activities, and sleep disturbance) were used to generate physical and mental health summary scores^36,71^. Higher domain scores indicate greater level of the construct and can be positive (e.g., physical function) or negative (e.g., fatigue).

##### EQ-5D-5L and Visual Analog Scale

Quality of life was measured using the interviewer-administered EuroQol EQ-5D-5L measure. The EQ-5D-5L asks respondents to assess five domains of health in the prior week including mobility, self-care, usual activities, pain, and anxiety/depression^38^. The data were summarized as a single index value using the United States EQ-5D-5L value sets^37,72^. Participants are also asked to rate their health in the preceding week on a 100-point visual analog scale (VAS) with 100 representing “best imaginable health state” and 0 representing the “worst imaginable health state.”

##### Patient Global Impression of Change

The self-reported Patient Global Impression of Change (PGIC) measure represents a participant’s opinion of the efficacy of an intervention and has been used in the context of pain disorders^41^ as well as depression, non-small cell lung cancer, and asthma^73^. Participants were asked to choose from a seven-point scale which phrase best represented their change with regard to activity limitations, symptoms, emotions and overall quality of life, related to Long COVID due to the study intervention. In a similar manner, they are asked the same question with a scale numbered zero to ten with zero representing “much better,” five representing “no change,” and ten representing “much worse.” A free response section was included below to further elaborate on symptom changes, if present.

##### World Health Organization Disability Assessment Schedule

The 12-item, interviewer-administered World Health Organization (WHO) Disability Assessment Schedule 2.0 (WHODAS 2.0) is a generic assessment instrument for health and disability covering six domains of function including cognition, mobility, self-care, interactions with others, life activities, and participation in community activities^39^. The simple scoring method according to the WHO manual for administration was utilized for analysis with higher scores signifying greater disability.

##### Duke Activity Status Index

The Duke Activity Status Index is a 12-item, patient-reported measure of functional capacity, demonstrated to correlate with peak oxygen uptake^42^. It gives a summary score from 0 to 58.2 where higher scores indicate higher functional capacity. Summary scores can be converted into estimates of peak oxygen uptake and maximal metabolic equivalent of tasks (METs).

##### Everyday Cognition Scale

The self-reported Everyday Cognition (ECog) 39-item scale was used to measure daily cognitive functioning^45^. The ECog was modified to assess participants’ current perception of neurocognitive status at the current timepoint compared to before they were ever diagnosed with COVID-19. A total score was calculated by summing the completed items and then dividing by the number of items completed. Higher scores indicate greater cognitive impairment. To utilize the instrument in the context of a clinical trial with frequent administration, we modified the time scale of the questionnaire, specifically assessing the change in participants’ cognitive functioning from before they ever tested positive for COVID-19 to the present day (in contrast to the original measure which compares cognitive functioning 10 years prior to the present day).

##### COMPASS-31

The COMPASS-31 is a 31-item, patient-reported scale that results in a composite score describing the degree of autonomic symptoms including secretomotor dysfunction, orthostatic intolerance, constipation, diarrhea, and more^47^. Higher composite scores indicate a greater degree of autonomic dysfunction. We modified the COMPASS-31 time scale to assess participants’ autonomic dysfunction over the prior 3 months (instead of 12 months as is included in the standard form) to accommodate the structure of the trial.

##### DSQ-PEM

We used a modified version of the DSQ-PEM to assess PEM^33^. In short, the DSQ-PEM assesses both frequency and severity of five post-exertional malaise symptoms in addition to five supplemental items to determine PEM duration, recovery, and exercise avoidance. Higher scores indicate greater burden of PEM symptoms. In order to use the measure in the context of the trial, we modified the time scale of the questionnaire, asking participants to assess PEM symptoms over the prior 3 months as opposed to the standard six-month time frame.

##### Other infection-associated conditions

We used standardized questionnaires to inquire about preceding diagnoses and current symptoms related to ME/CFS, hypermobile Ehlers-Danlos Syndrome, and endometriosis at participant screening visits. Participants with a prior diagnosis of ME/CFS preceding and not worsened by their subsequent development of Long COVID were excluded from participation according to pre-specified exclusion criteria. To assess if participants met criteria for ME/CFS after development of Long COVID symptoms, we utilized both the ME/CFS International Consensus Criteria^74^ and the Institute of Medicine (IOM) 2015 Diagnostic Criteria^32^ and assessed for severity of symptoms using the Bell CFIDS Disability Scale^75^. Hypermobile Ehlers-Danlos Syndrome was evaluated using the 2017 International Classification of the Ehlers-Danlos Syndrome criteria^76^, which includes a “Beighton score” to assess generalized joint hypermobility and includes diagnostic thresholds. To evaluate endometriosis, patients assigned female sex at birth completed the ENDOPAIN 4D Questionnaire^77^ which is divided into 21 items across 4 domains; total scores range from 0 to 94 with higher scores indicating greater burden of symptoms. Presence and history of mast cell activation syndrome (MCAS) was evaluated through Coastal Integrative Medicine’s “Mast Cell Activation Questionnaire”^78^, a 30-item assessment evaluating symptoms, medical history, and laboratory measurements; a total score above 8 but less than 14 indicates a pathological activation of mast cells, while a total score of 14 and more indicates a systemic mast cell mediator release syndrome and is considered clinically verified. The Mast Cell Activation Questionnaire was modified for ease of administration in the setting of an outpatient clinical trial.

#### Objective Clinical Assessments

##### Assessment of neurocognitive function

We used CNS Vital Signs^46^, a computer-based neurocognitive testing battery comprised of seven tests: verbal and visual memory, finger tapping, symbol digit coding, the Stroop Test, a test of shifting attention and the continuous performance test. The battery gives a summary neurocognition index (NCI) score averaging five domain scores (Composite Memory, Psychomotor Speed, Reaction Time, Complex Attention, and Cognitive Flexibility) and representing a global score of neurocognition.

##### Assessment of physical function

The six-minute walk test (6MWT) is a simple exercise testing procedure designed to assess physical ability through a six-minute, self-paced walk^43,44^. Measures of heart rate, oxygen saturation, blood pressure, and self-reported dyspnea and fatigue are performed before and after the test. Total distance walked in meters at each timepoint is reported.

##### Assessment of autonomic function

The active standing test is a non-invasive tool to assess orthostatic hypotension (OH) and postural orthostatic tachycardia syndrome (POTS). Procedures were similar to previously reported methods^48^. Blood pressure and heart rate measurements were obtained after 5 minutes of resting supine and 1, 3, 5, and 10 minutes of continuous standing. Abnormal active standing test results were defined as those with a decline of >20 mmHg in systolic or > 10 mmHg in diastolic blood pressure in at least two consecutive measurements, or those with an increase in heart rate > 30 bpm on two consecutive measurements^79^.

#### Laboratory Measurements

##### Clinical laboratory measurements

In addition to safety laboratory tests, C-reactive protein (CRP), erythrocyte sedimentation rate (ESR), fibrinogen, and D-dimer were measured in the clinical laboratory at San Francisco General Hospital.

##### Plasma cytokine levels

Cytokine levels were measured in plasma using the CorPlex Cytokine Panel (Quanterix). Plasma samples were diluted 4-fold in sample diluent buffer and assays were performed following the CorPlex manufacturer protocols. Each CorPlex Cytokine Panel kit was analyzed by the SP-X Imaging and Analysis System (Quanterix).

##### Anti-SARS-CoV-2 IgG antibody levels

Anti-SARS-CoV-2 IgG antibodies were measured in plasma samples using the previously described Simoa assays^80^. Antibody levels are presented as normalized average enzymes per bead (AEB), where the measured AEB value is normalized by calibrators obtained from serially diluting a plasma sample collected from a SARS-CoV-2 positive individual.

##### Plasma proteomic measurements

Plasma proteomic measurements were obtained from 1,034 unique proteins quantified in singlicate from cryopreserved plasma using the Olink® Reveal platform (Olink® Proteomics, Uppsala, Sweden), a highly sensitive and specific proximity extension assay (PEA)–based technology.^50^ For longitudinal analyses, multiple samples from the same donor were placed on the same plate to minimize inter-plate variability. Within each plate, samples were randomized using Olink® Analyze according to the manufacturer’s guidance. After PEA incubation and PCR amplification, DNA libraries were sequenced on an Illumina NovaSeq X Plus (Illumina Inc., San Diego, CA, USA) at the UCSF CAT Core using the *Olink_NovaSeqX_10B_8Lib_V3* recipe. Relative protein levels, expressed as NPX values (Normalized Protein eXpression; log₂ scale), were generated using ngs2counts (v5.1.0) and processed with NPX™ Map software (v1.3.0), including plate control normalization. All samples passed quality control, and 100% of data points met QC thresholds.

##### AER002 Pharmacokinetics

AER002 levels were measured via a validated serum LC-MS/MS method quantifying a unique CDR1-region signature peptide, developed based on a previously described method^31^.

A 50 µL serum aliquot was mixed with 750 µL of water/water saturated with ammonium sulfate (1:1, v/v), vortexed, and incubated at room temperature for ≥15 minutes. Samples were centrifuged at 15,000 rpm for 15 minutes at 5 °C, followed by another 15-minute incubation at room temperature. The supernatant was discarded, and 100 µL of denaturing buffer (8 M urea in 50 mM ammonium bicarbonate with 0.2 M DTT, 20:1, v/v) was added. Samples were incubated at 60 °C for 1 hour, vortexed, then alkylated by adding 20 µL of 500 mM iodoacetamide (final IAA concentration: 40 mM), followed by incubation in the dark at room temperature for 30 minutes. Excess IAA was quenched by adding 15 µL of 200 mM DTT. Next, 480 µL of 50 mM ammonium bicarbonate digestion buffer was added. Trypsin working solution was prepared by diluting 100 µL of 1 µg/µL trypsin stock in 900 µL buffer, and 50 µL of this solution was added to each sample. Internal standard (IS) solution (60 µL of 5,000 ng/mL) was added to all samples except double blanks. Samples were incubated at 37 °C for 6 hours, then digestion was quenched by adding 25 µL of water/TFA (9:1, v/v). The samples were centrifuged at 15,000 rpm for 15 minutes at 5 °C.

Supernatants were subjected to solid-phase extraction (SPE) for peptide purification prior to LC–MS/MS analysis. A 10 µL aliquot of each SPE-processed digest was injected into the LC–MS/MS system equipped with a Turbo V electrospray ionization (ESI) source operating in positive ion mode. Quantification of AER002 levels and their stable isotope-labeled internal standard, AER002 [13C6 15N4]FDDYALHWVR, was performed using multiple reaction monitoring (MRM) targeting a unique signature peptide derived from the CDR1 region. The ion source parameters were set as follows: ion spray voltage, 4500 V; source temperature, 500 °C; curtain gas, 30 psi; nebulizer gas (gas 1), 50 psi; heater gas (gas 2), 50 psi; and collision gas, 9 psi. MRM transitions monitored included m/z 441.2 → 530.4 for the unlabeled peptide and m/z 444.5 → 666.5 for the labeled internal standard.

#### Optional Procedures

##### Gut biopsy

Rectosigmoid tissue samples were collected via flexible sigmoidoscopy on an opt-in basis. Up to two collections occurred, once at baseline prior to infusion and another shortly after the 90-day follow-up time point. Tissue samples were fixed in fresh paraformaldehyde (PFA) followed by paraffin embedding approximately 24 hours after fixation or were cryopreserved at −180°C in fetal bovine serum (FBS) and 20% dimethyl sulfoxide (DMSO) according to previously described methods^15^.

The manual RNAscope 2.5 HD Duplex assay (Advanced Cell Diagnostics, catalog no. 322430) was used to identify SARS-CoV-2 single-stranded spike RNA [probe-V-nCoV2019-S (catalog no. 848561-C2)] and double-stranded orf1ab RNA [V-nCoV2019-orf1ab-sense (catalog no. 859151)] in situ. Paraffin-embedded tissue blocks were sectioned at 5 μm and mounted onto SuperFrost Plus slides, which were stored in a desiccant chamber prior to staining. Slides were baked in a dry air oven at 60°C for 1 hour, deparaffinized twice in 100% xylene (5 min), and dehydrated twice in 100% ethanol (1 min). To block endogenous peroxidase activity, slides were incubated with hydrogen peroxide for 10 minutes. Next, heat-induced epitope retrieval was performed for 18 minutes at 100°C using Target Retrieval Reagent (ACDBio). Protease digestion was accomplished by treatment with Protease Plus solution (ACDBio) for 30 min at 40°C. Hybridization was then performed with a duplex cocktail of probe-V-nCoV2019-orf1ab-sense (catalog no. 859151, 1:1 dilution) and probe-V-nCoV2019-S (catalog no. 848561-C2, 1:50 dilution) for 2 hours at 40°C. Following hybridization, amplifications one through ten, as well as detection of the red and green channels, were performed according to the manufacturer’s original protocol. Following detection of the green channel, slides were counterstained with Gill’s Hematoxylin I (StatLab, catalog no. HXGHE1LT, 1:2 dilution), blued with .02% ammonia water (Thermo Scientific Chemicals, catalog no. 1336-21-6), and mounted using VectaMount Permanent Mounting Medium (H-5600-60).

Images were captured using the Keyence BZ-X810. FFPE rectal tissues from pre-pandemic participants, as well as FFPE lung tissue from SARS-CoV-2 infected K18-hACE2 mice, were run as negative and positive controls, respectively. Sections from all FFPE blocks were also stained with a human housekeeping gene cocktail (POL2RA, PPIB) to assess block RNA integrity and assay performance.

We used the nCounter Host Immune Response RNA expression panel (nanoString) to determine differential expression of 750 human gene transcripts related to viral infection response and inflammation, as well as a custom SARS-CoV-2 RNA panel containing single- and double-stranded genes across the viral genome. Pre-COVID rectosigmoid tissue samples were used to identify false-positive signal and determine thresholds of positivity of SARS-CoV-2 transcript quantitation in the post-COVID samples. nCounter ROSALIND software was used to perform differential gene expression analyses between treatment groups and between baseline and post-infusion timepoints for each study arm. This approach selects the optimal subset of housekeeping probes for normalization using the geNorm algorithm, as implemented in the Bioconductor package. Differential expression between two groups of samples is calculated using the Generalized Linear Model (GLM) for count data assuming a negative binomial distribution and utilizes the raw data and estimates of noise and dispersion across all samples in the experiment to calculate fold-change and p-value for each gene. Prior to DEG analyses, ROSALIND was used to remove genes/probes from differential expression analysis if they are expressed near or at background levels and then to normalize raw data. First, geometric mean of the selected housekeeping genes was calculated for each sample followed by geometric mean calculations across all sample-specific geometric means to generate a global geometric mean. Next, the global geometric mean was divided by the sample-specific geometric mean for each sample to generate a normalization factor for each sample followed by raw counts multiplied by of each gene by its sample-specific normalization factor.

##### [18F]F-AraG PET Imaging

Participants were administered intravenously [18F]F-AraG (∼5 mCi) and PET-CT whole-body imaging was carried out for 20 minutes at approximately 50 minutes post injection. [18F]F-AraG was prepared as documented elsewhere^15^. Images were taken from the top of head to mid thighs. Routine urinalysis was performed 7 to 14 days after imaging to ensure proper excretion of [18F]F-AraG. Standardized uptake values (SUV) in various tissue regions of interest (ROI) from PET-CT data were determined using the OsiriX DICOM viewer software package (Pixmeo). ROI determination was performed in complex structures such as brain sections, heart wall, spleen, and gut wall using two-dimensional isometric ROIs as previously described^15^. For simpler structures such as the spinal cord, bone marrow, and whole lymph nodes, three-dimensional spherical VOIs were used). ROI were placed independently by two individuals blinded to the study group following ROI determination upon a subcohort and comparison to ensure consistency across reviewers. The non-parametric Mann-Whitney U testing (double sided with alpha level of 0.05) was used to compare SUV in cross sectional analyses and the Wilcoxon signed-rank test (two-sided with alpha level of 0.05) was used for paired intra-arm analyses before and after treatment. Strict adjustments for multiple comparisons were not used given the limited sample size as these were hypothesis-generating, exploratory analyses. All PET images and CT images were reviewed by board-certified radiologists for incidental abnormalities that would require clinical follow up.

##### Cardiopulmonary exercise testing

Cardiopulmonary exercise testing (CPET) procedures were performed according to our standard research protocol^81^ on an opt-in basis once between screening and infusion, and again shortly after the D90 follow-up time point to avoid biasing or reducing statistical power of the main study analyses. Participants were invited to complete symptom-limited, maximal-effort CPET on an upright cycle ergometer using a metabolic cart (MGC Diagnostics Corporation) to objectively measure exercise capacity by assessing oxygen consumption at peak exercise (peak VO2 relative to body weight, ml/kg/min and percent predicted by the Wasserman equations) and peak power output (Watts). Ramp protocols were determined targeting a 10-minute test based on measured maximal voluntary ventilation rounding down to the nearest 5 Watt/minute ramp protocol (minimum 10 Watts/minute); the same protocol was used for both baseline and follow-up tests. CPETs were interpreted blinded to treatment assignment.

#### Statistical Analysis

##### Primary and secondary outcomes

All randomized participants were included in the main analysis. Linear mixed effects models were used for the primary and secondary patient reported outcomes, with an indicator for randomization group, visit and a visit-by-randomization group interaction. Lab measurement outcomes were log transformed. The visit variable was categorical and corresponded to a study visits post infusion. Models were adjusted for baseline scores and included an unstructured covariance structure. If an outcome was measured at both a screening and baseline visit, an average baseline score was calculated and incorporated in the baseline adjusted models. Baseline adjusted mean scores were obtained and reported for each time point by margining the linear mixed effect models^82^ Baseline adjusted geometric means and percent differences are reported for log-transformed laboratory outcomes. A per protocol analysis was conducted excluding participants who had a confirmed or suspected reinfection before the primary endpoint. All statistical analyses were carried out in R (version 4.4.1)^83^. Additional information regarding the statistical analyses can be found in the Statistical Analysis Plan.

##### Exploratory outcomes

Responders were defined as individuals reporting PGIC improvements at D90 and D180 (the blinded period) of “Moderately Better” or greater. Baseline antibody levels were compared between AER002 and placebo groups, as well as responders and non-responders using Wilcoxon rank-sum test.

Plasma proteomics data were analyzed using linear mixed-effects models fit separately for each protein, with fixed effects for randomization group, visit, and a visit-by-randomization group interaction. A random intercept for each participant was included to account for within-participant correlation. The interaction term was used to assess evidence for differential protein expression. The estimates for the interaction term were used to generate volcano plots, and p values from the models were corrected for multiple testing using the Storey Q method^51^.

Exploratory gene set enrichment analyses were performed using the *clusterProfiler (version 4.16.0)*^84^ package. Proteins were mapped to Entrez Gene IDs, ranked by fold changes and enrichment was tested against the Gene Ontology Biological Process^85^ and Hallmark gene sets^52^.

AER002 PK was characterized using noncompartmental and population pharmacokinetic approaches using R (version 4.5) and NONMEM (ICON, plc, version 7.6), respectively. All lower limit of quantitation (LLOQ) values (i.e., < 20 mcg/ml) were set to zero. One, two, and three compartment models with first-order elimination were evaluated. Between participant variability was explored on model parameters, alongside assessment of additive, proportional, and combined residual error models. The final model was selected based on goodness-of-fit diagnostics, visual predictive checks, bootstrap estimates, and comparison of objective function values for nested models (−2LL and AIC criterion). Covariate analysis to evaluate impacts of sex, age, and race was carried out using stepwise covariate modeling^86^.

As part of a post-hoc exploratory analysis assessing how drug exposure and baseline antibody levels affected response, multiple logistic regression was performed using AER002 AUC (area under the concentration-time curve from noncompartmental analysis) and baseline anti-SARS-CoV-2 levels (high [≥median] vs low [<median] anti-S, anti-S1, anti-RBD, anti-N levels separately) as predictors of whether an individual self-identified as a responder.

## FOOTNOTES

## Acknowledgements

We acknowledge the study participants, their caregivers, and their medical providers. We also acknowledge the following for their contributions: Monika Deswal and Elnaz Eilkhani for regulatory support; the UCSF LIINC, RECOVER, and clinical trials teams especially Viva Tai, Khamal Anglin, Grace Anderson, Melissa Buitrago, Emily Conway, Avery Eun, and Megan Lew for participant referrals and coordination; the UCSF Investigational Drug Service, especially Scott Fields, for preparing and dispensing study product; the UCSF Parnassus Clinical Research Service (CRS), especially Josephine Liu, Ryan Gunay, Patricia Cannon, Beryl Abungan, and Allison Ingebretson for administering the study drug and monitoring participants; the UCSF Specimen Processing & Banking Subcore including Salman Mahboob and Billy Huang for overseeing biospecimen processing; the Carrington Lab (National Cancer Institute) for performing HLA typing that allowed selection of participants for T cell analyses; the Henrich lab, especially Lilian Grimbert and Belen Altamirano Poblano for assistance with the tissue-based assays; Danny Li and Marty Levkova-Clark for assistance in performing the cardiopulmonary exercise tests; Safety Monitoring Committee members Vincent Marconi, Jason Goldman, and Rasika Karnik; Molly Beam for guidance on CNS-Vital Signs neurocognitive testing; Lisa McCorkell and Hannah Davis from the Patient-Led Research Collaborative for advice in the design and implementation of the study; and Amy Proal for input on the manuscript.

## Funding

The conduct of the clinical trial was financially supported by the Patient-Led Research Fund, a project of Patient-Led Research Collaborative funded by Balvi. The optional gut biopsy and imaging procedures were funded by the PolyBio Research Foundation. Additional biological measurements were supported by funding from the National Institute for Neurologic Disorders and Stroke (NINDS; R01NS136197) and the PolyBio Research Foundation. This project was supported by the National Center for Advancing Translational Sciences, National Institutes of Health, through UCSF-CTSI Grant Number UL1 TR001872. Sequencing was performed at the UCSF CAT, supported by UCSF PBBR, RRP IMIA, and NIH 1S10OD028511-01 grants. The contents of this manuscript are solely the responsibility of the authors and do not necessarily represent the official views of the NIH.

## Conflicts of interest

MJP has received consulting fees from Gilead Sciences, AstraZeneca, BioVie, Apellis Pharmaceuticals, BioNTech, and TechImmune, travel support from Invivyd, and research support from Shionogi, BioVie, and ImmunityBio, outside the submitted work. YF reports a financial interest in Eugit Therapeutics, which is unrelated to the submitted work. MSD has received consulting fees from Merck. DG has received consulting fees from Gilead, Merck, and Viiv. SGD reports consulting for Enanta Pharmaceuticals and Pfizer.

## Author Contributions

Obtained funding: MJP, SGD Trial design: MJP, DVG, SGD

Trial recruitment and study activities: DR, MCW, EAF, RH, KBP

Imaging recruitment and analysis: DR, KAA, EdN, RRF, HVB, TJH

Gut biopsy recruitment and analysis: DR, AER, BL, MS, TJH

Other laboratory analysis: MR, MP, EJW, ZNS, LMH, DRW, YF, PWH, TJH, AND

Patient advisors: AS, SB

Provision of study drug: HT, JA, PB, RV

Data analysis: TD, DHTC, BL, BH, DVG, AND, TJH

Data interpretation: MJP, TD, DHTC, BH, JNM, MSD, PWH, DVG, AND, TJH, SGD

Drafted initial manuscript: MJP, DR, TD, MSD, LC, AND, TJH, SGD

Edited and approved manuscript: All authors

## Data Availability

The protocol and statistical analysis plan are available on clinicaltrials.gov (NCT05877508). The data that support the findings of this study are available from the corresponding author upon reasonable request.

## Notes

### Clinical Trial

NCT05877508

### Clinical Protocols

https://clinicaltrials.gov/study/NCT05877508

### Author Declarations

Institutional Review Board of the University of California, San Francisco gave ethical approval for this work.

