## Supplemental Materials for "A phase 2a double-blind, placebo-controlled randomized trial of the SARS-CoV-2-specific monoclonal antibody AER002 in people with Long COVID"

**Supplemental Table 1. Related adverse events (AEs).**

| | AER002 | | | | Placebo | | | |
|---|---|---|---|---|---|---|---|---|
| Adverse Event (AE) | Mild (no. AEs) | Moderate (no. AEs) | Participants (n = 24) | % | Mild (no. AEs) | Moderate (no. AEs) | Participants (n = 12) | % |
| **Cardiovascular** | | | | | | | | |
| Hypotension | 0 | 0 | 0 | 0.0 | 1 | 0 | 1 | 8.3 |
| **Dermatologic** | | | | | | | | |
| Localized rash | 0 | 1 | 1 | 4.2 | 0 | 0 | 0 | 0.0 |
| Localized itchiness | 0 | 0 | 0 | 0.0 | 0 | 1 | 1 | 8.3 |
| **ENT** | | | | | | | | |
| Cold sores | 0 | 1 | 1 | 4.2 | 1 | 1 | 2 | 16.7 |
| Sore throat | 0 | 0 | 0 | 0.0 | 1 | 0 | 1 | 8.3 |
| **Gastrointestinal** | | | | | | | | |
| Constipation | 0 | 0 | 0 | 0.0 | 1 | 0 | 1 | 8.3 |
| Diarrhea | 0 | 0 | 0 | 0.0 | 3 | 0 | 3 | 25.0 |
| **General/systemic** | | | | | | | | |
| Post-exertional malaise | 2 | 1 | 3 | 12.5 | 0 | 4 | 4 | 33.3 |
| Fatigue | 2 | 0 | 2 | 8.3 | 0 | 0 | 0 | 0.0 |
| Chills | 1 | 0 | 1 | 4.2 | 0 | 0 | 0 | 0.0 |
| Subjective fever | 1 | 0 | 1 | 4.2 | 0 | 0 | 0 | 0.0 |
| Pre-syncope | 0 | 0 | 0 | 0.0 | 1 | 0 | 1 | 8.3 |
| **Musculoskeletal** | | | | | | | | |
| Muscle pain/aches | 4 | 1 | 5 | 20.8 | 0 | 0 | 0 | 0.0 |
| Pain at infusion site | 1 | 0 | 1 | 4.2 | 1 | 0 | 1 | 8.3 |
| Muscle twitching | 1 | 0 | 1 | 4.2 | 0 | 0 | 0 | 0.0 |
| **Neurologic** | | | | | | | | |
| Headache | 1 | 1 | 2 | 8.3 | 0 | 1 | 1 | 8.3 |
| Trouble with balance/feeling unsteady | 1 | 0 | 1 | 4.2 | 0 | 1 | 1 | 8.3 |
| Dizziness | 0 | 1 | 1 | 4.2 | 0 | 0 | 0 | 0.0 |
| Head pressure | 0 | 1 | 1 | 4.2 | 0 | 0 | 0 | 0.0 |
| Lightheadedness | 0 | 1 | 1 | 4.2 | 0 | 0 | 0 | 0.0 |
| Burning sensation in feet | 0 | 0 | 0 | 0.0 | 1 | 0 | 1 | 8.3 |
| Tingling | 0 | 0 | 0 | 0.0 | 1 | 0 | 1 | 8.3 |
| **Ocular** | | | | | | | | |
| Blurry vision | 0 | 1 | 1 | 4.2 | 0 | 0 | 0 | 0.0 |
| **Total** | 14 | 9 | — | — | 11 | 8 | — | — |

### Supplemental Table 2. Unrelated adverse events (AEs).

| Adverse Event (AE) | AER002 Mild (no. AEs) | AER002 Moderate (no. AEs) | AER002 Severe (no. AEs) | AER002 Participants (n = 24) | AER002 % | Placebo Mild (no. AEs) | Placebo Moderate (no. AEs) | Placebo Severe (no. AEs) | Placebo Participants (n = 12) | Placebo % |
|---|---|---|---|---|---|---|---|---|---|---|
| **Cardiovascular** | | | | | | | | | | |
| Superficial thrombophlebitis | 1 | 0 | 0 | 1 | 4.2 | 0 | 0 | 0 | 0 | 0.0 |
| Hypertension | 0 | 0 | 0 | 0 | 0.0 | 1 | 0 | 0 | 1 | 8.3 |
| **Dermatologic** | | | | | | | | | | |
| Bruise | 2 | 0 | 0 | 2 | 8.3 | 1 | 0 | 0 | 1 | 8.3 |
| Skin pain | 0 | 1 | 0 | 1 | 4.2 | 0 | 0 | 0 | 0 | 0.0 |
| **ENT** | | | | | | | | | | |
| Congestion | 1 | 0 | 0 | 1 | 4.2 | 0 | 0 | 0 | 0 | 0.0 |
| Sinusitis | 0 | 1 | 0 | 1 | 4.2 | 0 | 0 | 0 | 0 | 0.0 |
| Bronchitis | 0 | 0 | 0 | 0 | 0.0 | 0 | 2 | 0 | 1 | 8.3 |
| Chronic rhinosinusitis | 0 | 0 | 0 | 0 | 0.0 | 0 | 1 | 0 | 1 | 8.3 |
| **Gastrointestinal** | | | | | | | | | | |
| Gastroenteritis | 1 | 2 | 0 | 2 | 8.3 | 1 | 0 | 0 | 1 | 8.3 |
| Diarrhea | 1 | 0 | 0 | 1 | 4.2 | 0 | 0 | 0 | 0 | 0.0 |
| **General/systemic** | | | | | | | | | | |
| Pre-syncope | 1 | 1 | 0 | 2 | 8.3 | 0 | 0 | 0 | 0 | 0.0 |
| Fatigue | 0 | 1 | 0 | 1 | 4.2 | 0 | 0 | 0 | 0 | 0.0 |
| **Genitourinary** | | | | | | | | | | |
| Dysuria | 1 | 0 | 0 | 1 | 4.2 | 0 | 0 | 0 | 0 | 0.0 |
| Kidney stone | 0 | 1 | 0 | 1 | 4.2 | 0 | 0 | 0 | 0 | 0.0 |
| Urinary tract infection | 0 | 1 | 0 | 1 | 4.2 | 0 | 0 | 0 | 0 | 0.0 |
| Flank pain | 0 | 0 | 0 | 0 | 0.0 | 1 | 0 | 0 | 1 | 8.3 |
| **Infectious** | | | | | | | | | | |
| Non-specific viral infection | 5 | 2 | 0 | 7 | 29.2 | 2 | 1 | 0 | 2 | 16.7 |
| SARS-CoV-2 infection - confirmed | 1 | 5 | 0 | 6 | 25.0 | 2 | 2 | 0 | 4 | 33.3 |
| SARS-CoV-2 infection - suspected[†] | 0 | 1 | 0 | 1 | 4.2 | 1 | 0 | 0 | 1 | 8.3 |
| Skin/soft tissue infection | 0 | 1 | 0 | 1 | 4.2 | 1 | 0 | 0 | 1 | 8.3 |
| Rebound COVID-19 | 0 | 0 | 0 | 0 | 0.0 | 0 | 1 | 0 | 1 | 8.3 |
| Small intestinal bacterial overgrowth | 0 | 0 | 0 | 0 | 0.0 | 0 | 1 | 0 | 1 | 8.3 |
| Suspected oral infection | 0 | 0 | 0 | 0 | 0.0 | 0 | 1 | 0 | 1 | 8.3 |
| **Laboratory abnormality** | | | | | | | | | | |
| Decreased fibrinogen | 3 | 0 | 0 | 2 | 8.3 | 0 | 0 | 0 | 0 | 0.0 |
| Hypoglycemia | 1 | 0 | 0 | 1 | 4.2 | 0 | 0 | 0 | 0 | 0.0 |
| **Musculoskeletal** | | | | | | | | | | |
| Fibromyalgia | 1 | 0 | 0 | 1 | 4.2 | 0 | 0 | 0 | 0 | 0.0 |
| Muscle pain/aches | 0 | 1 | 0 | 1 | 4.2 | 0 | 0 | 0 | 0 | 0.0 |
| Pain | 0 | 2 | 0 | 1 | 4.2 | 0 | 0 | 0 | 0 | 0.0 |
| Pelvic congestion syndrome | 0 | 1 | 0 | 1 | 4.2 | 0 | 0 | 0 | 0 | 0.0 |
| Gout flare | 0 | 0 | 0 | 0 | 0.0 | 0 | 1 | 0 | 1 | 8.3 |
| Rotator cuff tear | 0 | 0 | 0 | 0 | 0.0 | 0 | 1 | 0 | 1 | 8.3 |
| Torn meniscus | 0 | 0 | 0 | 0 | 0.0 | 0 | 1 | 0 | 1 | 8.3 |
| Torn patellar cartilage | 0 | 0 | 0 | 0 | 0.0 | 0 | 1 | 0 | 1 | 8.3 |
| Torn pulley ligament | 0 | 0 | 0 | 0 | 0.0 | 0 | 1 | 0 | 1 | 8.3 |
| **Neurologic** | | | | | | | | | | |
| Brain fog | 0 | 1 | 0 | 1 | 4.2 | 0 | 0 | 0 | 0 | 0.0 |
| Intractable chronic migraine with aura | 0 | 0 | 1 | 1 | 4.2 | 0 | 0 | 0 | 0 | 0.0 |
| Numbness | 0 | 1 | 0 | 1 | 4.2 | 0 | 0 | 0 | 0 | 0.0 |
| Abnormal brain MRI | 0 | 0 | 0 | 0 | 0.0 | 0 | 1 | 0 | 1 | 8.3 |
| **Ocular** | | | | | | | | | | |
| Keratoconjunctivitis sicca | 0 | 0 | 0 | 0 | 0.0 | 1 | 0 | 0 | 1 | 8.3 |
| Light sensitivity | 0 | 0 | 0 | 0 | 0.0 | 0 | 1 | 0 | 1 | 8.3 |
| **Total** | 19 | 23 | 1 | — | — | 11 | 16 | 0 | — | — |

[†] Had known exposure to SARS-CoV-2, but subsequently tested negative

**Supplemental Table 3. Pre-treatment values for continuous patient reported outcomes, performance measures, and laboratory measurements.**

| Patient Reported Outcome | AER002* | Placebo* | Mean difference | 95% CI | p-value |
|---|---|---|---|---|---|
| **PROMIS-29 PHSS** (T-score) | 39.7 | 38.0 | 1.7 | (-5.4, 8.9) | 0.62 |
| **PROMIS-29 MHSS** (T-score) | 37.5 | 37.6 | -0.1 | (-4.9, 4.6) | 0.96 |
| **QoL 100-point VAS** | 49.1 | 44.2 | 4.9 | (-9.0, 18.8) | 0.48 |
| **QoL EQ-5D-5L Index** | 0.48 | 0.49 | -0.01 | (-0.21, 0.18) | 0.90 |
| **DASI** | 36.5 | 28.6 | 7.9 | (-3.9, 19.7) | 0.18 |
| **COMPASS-31** | 20.2 | 28.5 | -8.3 | (-19.6, 2.9) | 0.14 |
| **WHO-DAS 2.0** | 17.3 | 18.8 | -1.5 | (-7.2, 4.2) | 0.60 |
| **ECog-39** | 2.29 | 2.49 | -0.20 | (-0.72, 0.30) | 0.42 |
| **6MWT** (meters) | 412 | 400 | 12 | (-76.9, 101) | 0.78 |
| **CNS-VS NCI** | 95.6 | 93.2 | 2.4 | (-4.6, 9.3) | 0.49 |
| **Laboratory measurements** | **AER002**** | **Placebo**** | **Percent difference** | **95% CI** | **p-value** |
| **CRP** (mg/mL) | 0.64 | 0.69 | -7.1 | (-58.8, 109.5) | 0.86 |
| **ESR** (mm/hr) | 9.52 | 8.50 | 12.1 | (-40.1, 109.8) | 0.71 |
| **D-Dimer** (mg/L FEU) | 0.29 | 0.33 | -13.2 | (-27.7, 4.16) | 0.12 |
| **Fibrinogen** (mg/dL) | 288 | 298 | -3.2 | (-16.7, 12.4) | 0.66 |

*Estimated marginal means **Estimated marginal geometric means

**Supplemental Table 4. Linear mixed effect model results for continuous patient reported outcomes, performance measures and laboratory measurements.**

| Patient Reported Outcome | Day | AER002* | Placebo* | Mean difference | 95% CI | p-value |
|---|---|---|---|---|---|---|
| **PROMIS-29 PHSS** (T-score) | 30 | 40.7 | 41.6 | -0.9 | (-5.8, 4.1) | 0.73 |
| | 90 | 40.9 | 44.9 | -4.0 | (-8.9, 1.0) | 0.12 |
| | 180 | 41.7 | 44.3 | -2.5 | (-7.5, 2.4) | 0.31 |
| | 360 | 42.6 | 46.6 | -4.0 | (-8.9, 0.96) | 0.11 |
| **PROMIS-29 MHSS** (T-score) | 30 | 40.3 | 42.7 | -2.4 | (-7.7, 3.0) | 0.38 |
| | 90 | 40.7 | 45.4 | -4.7 | (-10.0, 0.68) | 0.09 |
| | 180 | 41.3 | 43.8 | -2.6 | (-7.9, 2.8) | 0.34 |
| | 360 | 43.6 | 45.4 | -1.8 | (-7.1, 3.6) | 0.52 |
| **QoL 100-point VAS** | 30 | 54.4 | 55.7 | -1.3 | (-13.2, 10.6) | 0.83 |
| | 90 | 53.4 | 62.2 | -8.1 | (-20.7, 3.1) | 0.14 |
| | 180 | 54.8 | 61.1 | -6.3 | (-18.2, 5.6) | 0.29 |
| | 360 | 58.6 | 62.3 | -3.8 | (-15.6, 8.1) | 0.53 |
| **QoL EQ-5D-5L Index** | 30 | 0.58 | 0.56 | 0.02 | (-0.16, 0.19) | 0.85 |
| | 90 | 0.51 | 0.67 | -0.15 | (-0.32, 0.02) | 0.08 |
| | 180 | 0.60 | 0.64 | -0.03 | (-0.21, 0.14) | 0.69 |
| | 360 | 0.65 | 0.63 | 0.02 | (-0.15, 0.19) | 0.84 |
| **DASI** | 30 | 35.0 | 37.3 | -2.3 | (-10.1, 5.3) | 0.54 |
| | 90 | 35.8 | 39.9 | -4.1 | (-11.8, 3.6) | 0.30 |
| | 180 | 33.8 | 41.6 | -7.1 | (-15.4, 0.01) | 0.05 |
| | 360 | 39.8 | 43.3 | -3.5 | (-11.2,4.2) | 0.37 |
| **COMPASS-31** | 90 | 18.2 | 13.8 | 4.4 | (-3.9, 12.9) | 0.29 |
| | 180 | 21.3 | 19.2 | 2.1 | (-6.3, 10.6) | 0.62 |
| | 360 | 17.4 | 20.7 | -3.3 | (-11.7, 5.2) | 0.44 |
| **WHO-DAS 2.0** | 90 | 16.8 | 12.9 | 3.9 | (-0.95, 8.7) | 0.11 |
| | 180 | 14.9 | 14.1 | 0.8 | (-4.0, 5.6) | 0.75 |
| | 360 | 12.4 | 11.7 | 0.6 | (-4.2, 5.5) | 0.78 |
| **ECog-39** | 30 | 2.18 | 2.01 | 0.17 | (-0.18, 0.53) | 0.34 |
| | 90 | 2.09 | 1.93 | 0.16 | (-0.19, 0.51) | 0.37 |
| | 180 | 2.04 | 2.11 | -0.07 | (-0.42, 0.29) | 0.71 |
| | 360 | 1.94 | 1.99 | -0.05 | (-0.41, 0.30) | 0.76 |
| **6MWT** (meters) | 30 | 422 | 408 | 14.0 | (-34.8, 63.3) | 0.57 |
| | 90 | 424 | 455 | -31.0 | (-80.0, 18.2) | 0.21 |
| | 180 | 415 | 450 | -35.0 | (-84.2, 14.3) | 0.16 |
| | 360 | 430 | 474 | -44.0 | (-96.0, 8.2) | 0.10 |
| **CNS-VS NCI** | 90 | 99.3 | 100.7 | -1.4 | (-6.03, 3.2) | 0.55 |
| | 180 | 97.7 | 103.2 | -5.6 | (-10.2, 0.94) | 0.02 |
| | 360 | 102.8 | 102.9 | -0.1 | (-4.8, 4.6) | 0.97 |

| Laboratory Measurements | Day | AER002** | Placebo** | Percentage Difference | 95% CI | p-value |
|---|---|---|---|---|---|---|
| **CRP** (mg/mL) | 30 | 0.71 | 0.76 | -6.6 | (-55, 93) | 0.85 |
| | 90 | 0.82 | 0.72 | 14.1 | (-45, 136) | 0.72 |
| | 180 | 0.80 | 0.98 | -18.5 | (-61, 69) | 0.58 |
| | 360 | 0.76 | 1.50 | -49.3 | (-77, 14) | 0.10 |
| **ESR** (mm/hr) | 30 | 8.3 | 8.1 | 3.1 | (-37, 69) | 0.90 |
| | 90 | 9.0 | 9.5 | -5.5 | (-42, 55) | 0.82 |
| | 180 | 8.4 | 7.6 | 10.1 | (-33, 80) | 0.70 |
| | 360 | 7.4 | 9.5 | -21.5 | (-53, 32) | 0.36 |
| **D-Dimer** (mg/L FEU) | 30 | 0.31 | 0.32 | -4.7 | (-25, 21) | 0.69 |
| | 90 | 0.30 | 0.31 | -3.7 | (-24, 22) | 0.75 |
| | 180 | 0.32 | 0.38 | -17.5 | (-35, 5) | 0.11 |
| | 360 | 0.31 | 0.34 | -9.8 | (-29, 16) | 0.42 |
| **Fibrinogen** (mg/dL) | 30 | 292 | 289 | 0.9 | (-9.5, 12.5) | 0.87 |
| | 90 | 308 | 293 | 5.2 | (-5.6, 17.3) | 0.36 |
| | 180 | 309 | 306 | 1.1 | (-9.6, 13.0) | 0.85 |
| | 360 | 289 | 309 | -3.5 | (-14.4, 8.7) | 0.55 |

*Estimated marginal means **Estimated marginal geometric means. Linear mixed effect models include visit terms for D30 (where measured), D90, D180 and D360.

**Supplemental Table 5. AER002 population pharmacokinetic (PK) model parameters.**

| Parameter[1,2] | Estimate | 90% C.I.[3] |
|---|---|---|
| $\theta_{CL}$ | 0.0655 | 0.0530 – 0.0863 |
| $\theta_{V\ Central}$ | 3.95 | 2.78 – 5.62 |
| $\theta_{Q}$ | 13.6 | 0.392 – 28.1 |
| $\theta_{V\ Peripheral}$ | 3.18 | 1.93 – 5.38 |
| $\theta_{Male\ on\ V\ Peripheral}$ [4] | 1.96 | 1.27 – 2.88 |
| $\Omega_{CL}$ | 0.354 | 0.049 |
| $\Omega_{V\ Peripheral}$ | 0.640 | 0.246 – 1.08 |
| $\sigma_{Proportional\ Error}$ | 0.228 | 0.122 – 0.285 |
| $\sigma_{Additive\ Error}$ | 12.1 | 5.14 – 20.4 |

1. $\theta$'s are the typical population estimates, $\Omega$'s are the estimates for between subject variability and $\sigma$'s are the estimates for residual unexplained variability.
2. Clearance (CL) in L/day, Volume of Distribution (V) in L/kg, Inter-compartmental CL (Q) in L/day
3. C.I. is the confidence interval from bootstrap
4. Covariate expressed in the multiplicative effect on peripheral volume of distribution

**Supplemental Table 6. Per protocol linear mixed effect model results for continuous patient reported outcomes, performance measures and laboratory measurements.**

| **Patient Reported Outcome** | **Day** | **AER002*** | **Placebo*** | **Mean difference** | **95% CI** | **p-value** |
|---|---|---|---|---|---|---|
| **PROMIS-29 PHSS** (T-score) | 90 | 40.8 | 44.1 | -3.3 | (-8.5, 1.7) | 0.18 |
| **PROMIS-29 MHSS** (T-score) | 90 | 41.5 | 43.1 | -1.6 | (-6.0, 2.7) | 0.45 |
| **QoL 100-point VAS** | 90 | 53.4 | 58.9 | -5.5 | (-16.3, 5.3) | 0.31 |
| **QoL EQ-5D-5L Index** | 90 | 0.55 | 0.63 | -0.08 | (-0.3, 0.1) | 0.34 |
| **DASI** | 90 | 36.3 | 38.8 | -2.5 | (-10.4, 5.5) | 0.53 |
| **COMPASS-31** | 90 | 17.8 | 15.4 | 2.4 | (-5.9, 10.9) | 0.55 |
| **WHO-DAS 2.0** | 90 | 16.9 | 13.4 | 3.5 | (-0.01, 7.1) | 0.05 |
| **ECog-39** | 90 | 2.04 | 1.94 | 0.1 | (-0.24, 0.45) | 0.53 |
| **6MWT** | 90 | 419 | 453 | -34 | (-83.4, 17.1) | 0.19 |
| **CNS-VS NCI score** | 90 | 101 | 101.2 | -0.2 | (-5.3, 4.9) | 0.94 |
| **Laboratory measurements** | **Day** | **AER002**** | **Placebo**** | **Percentage difference** | **95% CI** | **p-value** |
| **CRP** (mg/mL) | 90 | 0.78 | 0.63 | 22.6 | (-42, 156) | 0.58 |
| **ESR** (mm/hr) | 90 | 8.3 | 9.1 | -8.5 | (-43.3, 48.2) | 0.71 |
| **D-Dimer** (mg/L FEU) | 90 | 0.29 | 0.30 | -0.9 | (-9.7, 8.7) | 0.83 |
| **Fibrinogen** (mg/dL) | 90 | 306 | 288 | 6.1 | (-4.9, 18.4) | 0.28 |

*Estimated marginal means **Estimated marginal geometric means. Per protocol analyses exclude those with either a confirmed or suspected reinfection before the primary endpoint (D90)

**Supplemental Table 7. Optional Gut Biopsy RNAScope Results**

| PID | Treatment | Sex | Age (yrs) | Reinfection between biopsies | Timepoint (baseline vs. post-infusion) | Days since initial infection | Days since most recent infection | Spike RNA (SS) | orf1ab RNA (DS) |
|---|---|---|---|---|---|---|---|---|---|
| **1** | Placebo | Male | 70-79 | - | Baseline | 1315 | 1315 | - | - |
| | | | | | Post-infusion | 1434 | 1434 | - | - |
| **2** | Placebo | Female | 50-59 | - | Baseline | 1163 | 1163 | - | - |
| | | | | | Post-infusion | 1254 | 1254 | - | - |
| **3** | AER002 | Male | 50-59 | + | Baseline | 1093 | 1093 | - | - |
| | | | | | Post-infusion | 1198 | 21 | + | + |
| **4** | AER002 | Male | 50-59 | - | Baseline | 255 | 255 | - | - |
| | | | | | Post-infusion | 365 | 365 | - | - |
| **5** | AER002 | Male | 40-49 | + | Baseline | 515 | 515 | - | - |
| | | | | | Post-infusion | 620 | 28 | + | - |
| **6** | AER002 | Female | 30-39 | - | Baseline | 485 | 485 | - | - |
| | | | | | Post-infusion | 590 | 590 | - | - |
| **7** | AER002 | Female | 40-49 | - | Baseline | 752 | 318 | - | - |
| | | | | | Post-infusion | 871 | 437 | - | - |
| **8** | AER002 | Male | 30-39 | - | Baseline | 721 | 721 | - | - |
| | | | | | Post-infusion | 847 | 847 | - | - |
| **9** | AER002 | Male | 60-69 | - | Baseline | 559 | 559 | - | - |
| | | | | | Post-infusion | 671 | 671 | - | - |
| **10** | AER002 | Female | 40-49 | - | Baseline | 515 | 515 | - | - |
| | | | | | Post-infusion | 627 | 627 | - | - |
| **11** | AER002 | Male | 30-39 | - | Baseline | 448 | 448 | - | - |
| | | | | | Post-infusion | 546 | 546 | - | - |
| **12** | Placebo | Female | 40-49 | + | Baseline | 545 | 123 | - | - |
| | | | | | Post-infusion | 650 | 57 | - | - |
| **13** | Placebo | Male | 40-49 | - | Baseline | 811 | 811 | - | - |
| | | | | | Post-infusion | 930 | 930 | - | - |
| **14** | AER002 | Male | 40-49 | + | Baseline | 992 | 992 | - | - |
| | | | | | Post-infusion | 1111 | 21 | + | + |
| **15** | AER002 | Male | 50-59 | - | Baseline | 1238 | 718 | + | - |
| | | | | | Post-infusion | 1378 | 858 | - | - |
| **16** | AER002 | Female | 30-39 | - | Baseline | 729 | 396 | - | - |
| | | | | | Post-infusion | 833 | 500 | - | - |

| | | | | | | | | | |
|---|---|---|---|---|---|---|---|---|---|
| **17** | Placebo | Female | 40-49 | + | Baseline | 1093 | 1093 | - | - |
| | | | | | Post-infusion | 1288 | 18 | - | - |
| **18*** | AER002 | Female | 40-49 | - | Baseline | 1287 | 112 | - | - |

* Declined post biopsy

**Supplemental Table 8. Complete inclusion and exclusion criteria.**

| **Inclusion Criteria** |
|---|
| 1. Male, female, or transgender ≥18 years of age at Screening. |
| 2. Enrolled or willing to enroll and complete at least 1 visit in the UCSF Long-term Impact of Infection with Novel Coronavirus (LIINC) study. Any adult who has been infected with SARS-CoV-2 or has ever received or is eligible to receive a SARS-CoV-2 vaccination, does not have chronic anemia, and who is able to provide written informed consent, is eligible to participate in LIINC. |
| 3. History of confirmed acute SARS-CoV-2 infection, whose initial infection meets criteria as outlined below:<br>a. Report of a positive nucleic acid amplification test (NAAT) and/or a positive SARS-Cov-2 antigen rapid diagnostic test (RDT). Written proof of the test will be requested but is not required as long as the participant attests to the positive test.<br>AND<br>b. Long COVID (see below) attributed to SARS-CoV-2 infection with a variant against which AER002 is known to have neutralizing activity. In cases in which the variant is not known from prior viral sequencing (most cases), the SARS-CoV-2 infection to which Long COVID is attributed should have been dated prior to August 15, 2022.<br><br>*Note: For individuals who may be chronically infected (with persistent viral shedding from nasopharynx), a genotype is required to confirm that they have a AER002-susceptible variant.* |
| 4. Clinical evidence of Long COVID, as confirmed by the investigator's assessment.<br>a. At least two symptoms (see list) that are new or worsened since the time of SARS-CoV-2 infection, not known to be attributable to another cause upon assessment by the PI. At least two symptoms from those listed here must be present: systemic symptoms (e.g., fatigue, chills, post-exertional malaise), neurocognitive symptoms (e.g., trouble with memory/concentration ("brain fog"), headache, dysautonomia/postural orthostatic tachycardia syndrome, dizziness, unsteadiness, neuropathy, sleep disturbance), cardiopulmonary symptoms (e.g., chest pain, palpitations, shortness of breath, cough, fainting spells), musculoskeletal symptoms (e.g., muscle aches, joint pain), gastrointestinal symptoms (e.g., nausea, diarrhea). Although other symptoms (e.g., skin rash, hair loss, mental health symptoms, trouble with smell/taste, genitourinary symptoms) will be recorded and tracked, at least two core symptoms listed above must be present. Note: the two symptoms can be from within the same category (for example, brain fog and headache).<br>AND<br>b. Symptoms must have been present for at least 60 days prior to screening. Symptoms that wax and wane must have been initially present at least 60 days prior to screening<br>AND<br>c. Symptoms must be reported to be at least somewhat bothersome and to have an impact on quality of life and/or everyday functioning. |
| 5. Long COVID attributed to a SARS-CoV-2 infection prior to August 15, 2022. Note: While individuals re-infected with SARS-CoV-2 after August 15, 2022 will not be excluded, the SARS-CoV-2 infection after which Long COVID symptoms began must pre-date August 15, 2022. |
| 6. Not currently hospitalized. |
| 7. Body mass index (BMI) 18 to 50 kilograms/meter squared (kg/m2), inclusive, at the time of screening. |

| |
|---|
| 8. In otherwise stable health, as assessed by the investigator within 28 days prior to Screen, based on medical history, physical assessment, laboratory findings, and vital signs. |
| 9. Participants who are of childbearing potential (CBP) and male participants with sexual partner(s) who are females of CBP must agree to use adequate contraception from study consent through 360 days after dosing. Adequate contraception is defined as using hormonal contraceptives or an intrauterine device combined with at least 1 of the following forms of contraception: a diaphragm, a cervical cap, or a condom. Total abstinence from vaginal intercourse, in accordance with the lifestyle of the participant, is also acceptable. |
| 10. Willingness and ability to comply with the study protocol. This includes reliable transportation and sufficient time to attend all visits. |
| 11. Written informed consent (and assent when applicable) obtained from subject or subject's legal representative and ability for subject to comply with the requirements of the study. |

| **Exclusion Criteria** |
|---|
| 1. Current acute SARS-CoV-2 infection at the time of screening. |
| 2. Long COVID attributed to a SARS-CoV-2 infection after August 15, 2022. |
| 3. Previously received treatment or prophylaxis with a SARS-CoV-2-specific mAb, or plan to receive such treatment before exiting the study. |
| 4. Previously received COVID-19 convalescent plasma treatment within 60 days prior to planned Day 0 or plan to receive such treatment before exiting the study. |
| 5. Plans to receive any investigational or approved vaccine or booster for SARS-CoV-2 within 60 days prior to planned Day 0 or before Day 30 following planned Day 0. |
| 6. Active cardiovascular disease, defined as known prior:<br>a. Myocardial infarction within 90 days of screening<br>OR<br>b. Coronary artery bypass within 90 days of screening<br>OR<br>c. Current heart failure with reduced ejection fraction (<45%)<br>OR<br>d. Current pulmonary arterial hypertension. |
| 7. Known stroke within 3 months prior to planned Day 0. |
| 8. Known active bacterial, fungal, viral, or other infection besides SARS-CoV-2 requiring treatment within the 28 days prior to planned Day 0 and meeting criteria for systemic involvement upon review by the PI. Note: Mild or limited infections such as uncomplicated urinary tract or yeast infections, sexually transmitted infections, and mild dermatophyte infections may be reviewed with the medical monitor but are not exclusionary. |
| 9. Major surgery within 6 months prior to planned Day 0 or planned major surgery during the first 180 days following planned Day 0. |
| 10. History of unplanned hospitalization for >24 hours within 28 days prior to Screening. |
| 11. Active Hepatitis B (Hep B) infection (defined as Hep B surface antigen (sAg) positive).<br><br>*Note: A known positive Hep B core antibody (cAb) in the absence of positive sAg is not considered exclusionary.* |
| 12. Active Hepatitis C (Hep C) infection (defined as Hep C Ab positive or indeterminate with detectable Hep C RNA).<br><br>*Note: Those with cured Hep C (Ab positive or indeterminate but negative Hep C RNA) will remain eligible.* |

| |
|---|
| 13. HIV infection that is known to be unstable (two or more consecutive plasma HIV RNA values >48 copies/mL in the 6 months prior to screen) or uncontrolled (not on antiretroviral therapy (ART)). In addition, people with HIV who have a current CD4+ T cell count < 200 cells/uL, a history of AIDS defining illness without immune reconstitution, or clinical manifestations of symptomatic HIV will be excluded. |
| 14. Severe coagulopathy that would prevent an infusion (international normalized ratio ((INR) >2.0, history of hemophilia). |
| 15. Severe anemia (hemoglobin <9 grams/deciliter (g/dL)). |
| 16. Moderate or severe immunocompromise, according to the current NIH COVID-19 Treatment Guidelines as of March 6, 2023. This includes the following:<br>a. Receiving active treatment for solid tumor or hematologic malignancy, including use of systemic chemotherapy for treatment of cancer within the year prior to screening,<br>b. Prior solid-organ transplant with active immunosuppressive therapy<br>c. CAR-T cell therapy or hematopoietic cell transplant, on immunosuppressive therapy or transplant within the prior 2 years,<br>d. Primary immunodeficiency syndromes,<br>e. Advanced or untreated HIV infection (see above),<br>f. Or on active high-dose corticosteroids (i.e., >= 20mg prednisone or equivalent daily per day for >= 2 weeks). |
| 17. Known prior diagnosis of myalgic encephalomyelitis/chronic fatigue syndrome (ME/CFS), preceding and not related to SARS-CoV-2 infection and not worsened since SARS-CoV-2 infection. |
| 18. Known prior diagnosis of dysautonomia, preceding and not related to SARS-CoV-2 infection and not worsened since SARS-CoV-2 infection. |
| 19. History of anaphylaxis or hypersensitivity upon receiving IV antibody infusions in the past. |
| 20. Known allergy to any components used in the formulation of the intervention. |
| 21. History of anaphylaxis or similar significant allergic reaction to prescription or non-prescription drugs or food products. Similarly, presence of severe atopic conditions as assessed by the PI represents significant risk for allergic reaction. |
| 22. Pregnant, breastfeeding, or unwilling to practice birth control abide by the contraception requirements outlined in the inclusion criteria. |
| 23. Participation in a clinical trial with receipt of an investigational product within 28 days or 5 half-lives (whichever is longer) prior to planned Day 0. For PET tracers specifically, which have a short half-life and are not biologically active outside this window, elapse of 5 half-lives prior to the planned D0 infusion is sufficient. |
| 24. Current alcohol or illicit drug use as determined by the investigator to preclude participation. |
| 25. Presence of a condition or abnormality that in the opinion of the Investigator would compromise the safety of the patient or the quality of the data. |
| 26. Study site personnel directly affiliated with the study or family of directly involved personnel. |
| 27. History of cytokine release syndrome (CRS) secondary to infection and/or medication. |

**Supplemental Table 9. Descriptions of pre-specified primary and secondary outcomes**

| Primary Outcome | Day | Description |
|---|---|---|
| Patient-Reported Outcomes Measurement Information System (PROMIS)-29 Physical Health Summary Score (PROMIS-29 PHSS) | 90 | This measure will evaluate whether there is a difference between treatment with AER002 versus placebo in baseline adjusted mean PROMIS-29 Physical Health Summary Score at Day 90 post-infusion. PROMIS-29 is a validated scale assessing physical function, anxiety, depression, fatigue, sleep disturbance, ability to participate in social activities, and pain. Each domain is scored on a 5-point scale (without any difficulty, with a little difficulty, with some difficulty, with much difficulty, unable to do). A T-score is calculated from each individual domain. A T score of 50 represents the mean for US general adult population, and 10 is the standard deviation. A lower T score indicates worse physical health. |

| Category | Secondary Outcome | Day | Description |
|---|---|---|---|
| *Physical health and quality of life* | Patient-Reported Outcomes Measurement Information System (PROMIS)-29 Physical Health Summary Score (PROMIS-29 PHSS) | 30, 180 | This measure will evaluate whether there is a difference between treatment with AER002 versus placebo in baseline adjusted mean PROMIS-29 Physical Health Summary Score at Day 30 and Day 180 post-infusion. PROMIS-29 is a validated scale assessing physical function, anxiety, depression, fatigue, sleep disturbance, ability to participate in social activities, and pain. Each domain is scored on a 5-point scale (without any difficulty, with a little difficulty, with some difficulty, with much difficulty, unable to do). A T-score is calculated from each individual domain. A T score of 50 represents the mean for US general adult population, and 10 is the standard deviation. A lower T score indicates worse physical health. |
| *Physical health and quality of life* | Patient-Reported Outcomes Measurement Information System (PROMIS)-29 Mental Health Summary | 90 | This measure will evaluate whether there is a difference between treatment with AER002 versus placebo in baseline adjusted mean PROMIS-29 Mental Health Summary Score at Day 90 post-infusion. PROMIS-29 is a validated scale |

| | | | |
|---|---|---|---|
| | Score (PROMIS-29 MHSS) | | assessing physical function, anxiety, depression, fatigue, sleep disturbance, ability to participate in social activities, and pain. Each domain is scored on a 5-point scale (without any difficulty, with a little difficulty, with some difficulty, with much difficulty, unable to do). A T-score is calculated from each individual domain. A T score of 50 represents the mean for US general adult population, and 10 is the standard deviation. A lower T score indicates worse mental health. |
| *Physical health and quality of life* | Patient-Reported Outcomes Measurement Information System (PROMIS)-29 Mental Health Summary Score (PROMIS-29 MHSS) | 30, 180 | This measure will evaluate whether there is a difference between treatment with AER002 versus placebo in baseline adjusted mean PROMIS-29 Mental Health Summary Score at Day 30 and Day 180 post-infusion. PROMIS-29 is a validated scale assessing physical function, anxiety, depression, fatigue, sleep disturbance, ability to participate in social activities, and pain. Each domain is scored on a 5-point scale (without any difficulty, with a little difficulty, with some difficulty, with much difficulty, unable to do). A T-score is calculated from each individual domain. A T score of 50 represents the mean for US general adult population, and 10 is the standard deviation. A lower T score indicates worse mental health. |
| *Physical health and quality of life* | Quality of Life (Global Health Score) 100-point Visual-Analogue Scale | 90 | This measure will evaluate whether there is a difference between treatment with AER002 versus placebo in the baseline adjusted mean Quality of Life 100-point Visual-Analogue-Scale at Day 90 post-infusion. 0 represents the worst health a person can imagine and 100 represents the best health a person can imagine. |
| *Physical health and quality of life* | Quality of Life (5-Item EuroQol EQ-5D-5L) Index Value Score | 90 | This measure will evaluate whether there is a difference between treatment with AER002 versus placebo in baseline adjusted mean Quality of Life (5-Item EuroQol EQ- |

| | | | |
|---|---|---|---|
| | | | 5D-5L) Index Value Score at Day 90 post-infusion. 5-Item EuroQol EQ-5D-5L questions assess pain/difficulty in day-to-day activities over the past week. The 5-Item EuroQol EQ-5D-5L produces a score that typically ranges from 0 - 1, with a higher score indicating better quality of life. |
| *Physical health and quality of life* | World Health Organization Disability Assessment Schedule 2.0 (WHO-DAS 2.0) | 90 | This measure will evaluate whether there is a difference between treatment with AER002 versus placebo in baseline adjusted mean WHO-DAS 2.0 score at Day 90 post-infusion. The World Health Organization Disability Assessment Schedule 2.0 questionnaire asks about difficulties due to health conditions. Health conditions include diseases or illnesses, other health problems that may be short or long lasting, injuries, mental or emotional problems, and problems with alcohol or drugs. The range is scored from 0-48, with a higher score indicating a higher level of disability. |
| *Overall impression of change* | Patient Global Impression of Change (PGIC) Scale | 90 | This measure will evaluate whether there is a difference between treatment with AER002 versus placebo on the Patient Global Impression of Change (PGIC) scale at Day 90 post-infusion. The self-reported PGIC reflects a patient's belief about the efficacy of treatment. We used a modified PGIC scale which has been used to study pain syndromes and has been employed in other Long COVID clinical trials. It is a common data element developed by the National Institutes of Mental Health. The PGIC ranges from 0 (Much better) to 10 (Much Worse). A score of 5 indicates no change. |
| *Cardiopulmonary symptoms* | Duke Activity Status Index (DASI) | 90 | This measure will evaluate whether there is a difference between treatment with AER002 versus placebo in baseline adjusted mean DASI at Day 90 post-infusion. The Duke Activity Status Index is a patient-reported estimate of functional capacity, maximal oxygen |

| | | | |
|---|---|---|---|
| | | | consumption (VO2 max) and maximum metabolic equivalent of tasks (METs). The DASI questionnaire produces a score between 0 and 58.2 points, which is linearly correlated with a patient's VO2 max and METs, as measured from cardiopulmonary exercise testing (CPET). It inquires about a person's ability to perform self-care, walk, climb stairs, run, do house and yard work, engage in sexual intercourse, and perform moderate recreational activities. A higher score indicates higher functional capacity. |
| *Cardiopulmonary symptoms* | 6 Minute Walking Test (6MWT) | 90 | This measure will evaluate whether there is a difference between treatment with AER002 versus placebo in baseline adjusted mean distance walked on the 6MWT at Day 90 post-infusion. The 6MWT requires an individual to walk at their normal pace for 6 minutes on a marked track (for example, a hallway). Vital signs are assessed, and the total distance covered (in meters) is the primary outcome of interest. |
| *Neurocognitive symptoms and performance* | Everyday Cognition Form (ECog-39) | 90 | This measure will evaluate whether there is a difference between treatment with AER002 versus placebo in baseline adjusted mean ECog-39 score at Day 90 post-infusion. The ECog-39 is an instrument that measures the decline in everyday cognitive and functional abilities that map to six cognitive domains, adapted specifically to describe change in abilities since having COVID. A summary ECog-39 score is calculated scored with a range of 1-4, with a higher score indicating greater cognitive impairment. |
| *Neurocognitive symptoms and performance* | Neurocognition Index (NCI) standard score from the CNS-VS | 90 | This measure will evaluate whether there is a difference between treatment with AER002 versus placebo in baseline adjusted mean NCI standard score from the CNS-VS at Day 90 post-infusion. The CNS Vital Signs is a computer-based |

| | | | |
|---|---|---|---|
| | | | neurocognitive assessment comprised of seven tests: verbal and visual memory, finger tapping, symbol digit coding, the Stroop Test, a test of shifting attention and the continuous performance test. The battery gives a summary neurocognition index (NCI) score averaging five domain scores (Composite Memory, Psychomotor Speed, Reaction Time, Complex Attention, and Cognitive Flexibility) and representing a global score of neurocognition. NCI scores are normalized scores (mean 100, standard deviation 15) that are age matched relative to other people in a normative sample. A higher score indicates better cognitive function. |
| *Autonomic symptoms and performance* | Active Stand Test | 90 | The active standing test is a non-invasive tool to assess orthostatic hypotension (OH) and postural orthostatic tachycardia syndrome (POTS). In short, blood pressure and heart rate measurements were obtained after 5 minutes of resting supine and 1, 3, 5, and 10 minutes of continuous standing. Abnormal active standing test results were defined as those with a decline of >20 mmHg in systolic or > 10 mmHg in diastolic blood pressure in at least two consecutive measurements, or those with an increase in heart rate > 30 bpm on two consecutive measurements. |
| *Autonomic symptoms and performance* | Composite Autonomic Symptom Score-31 (COMPASS-31) | 90 | This measure will evaluate whether there is a difference between treatment with AER002 versus placebo in baseline adjusted mean COMPASS-31 score at Day 90 post-infusion. COMPASS-31 asks 31 questions related to autonomic dysfunction. The answer to each question generates a numeric score for the question, which is then summed at the end of the questionnaire. A total score out of 100 is generated summarizing orthostatic intolerance, vasomotor, secretomotor, gastrointestinal, |

| | | | |
|---|---|---|---|
| | | | urinary, pupillomotor, temperature intolerance, and sexual impairment. The total score ranges from 0 to 100 and a higher score indicates more severe autonomic dysfunction. |
| *Laboratory measurements* | C-Reactive Protein (CRP) | 90 | This measure will evaluate whether there is a difference between treatment with AER002 versus placebo in baseline adjusted mean CRP concentration (mg/L) at Day 90 post-infusion. |
| *Laboratory measurements* | Erythrocyte Sedimentation Rate (ESR) | 90 | This measure will evaluate whether there is a difference between treatment with AER002 versus placebo in baseline adjusted mean ESR at Day 90 post-infusion. |
| *Laboratory measurements* | D-Dimer | 90 | This measure will evaluate whether there is a difference between treatment with AER002 versus placebo in baseline adjusted mean D-Dimer at Day 90 post-infusion. |
| *Laboratory measurements* | Fibrinogen | 90 | This measure will evaluate whether there is a difference between treatment with AER002 versus placebo in baseline adjusted mean fibrinogen concentration (mg/dL) at Day 90 post-infusion. |

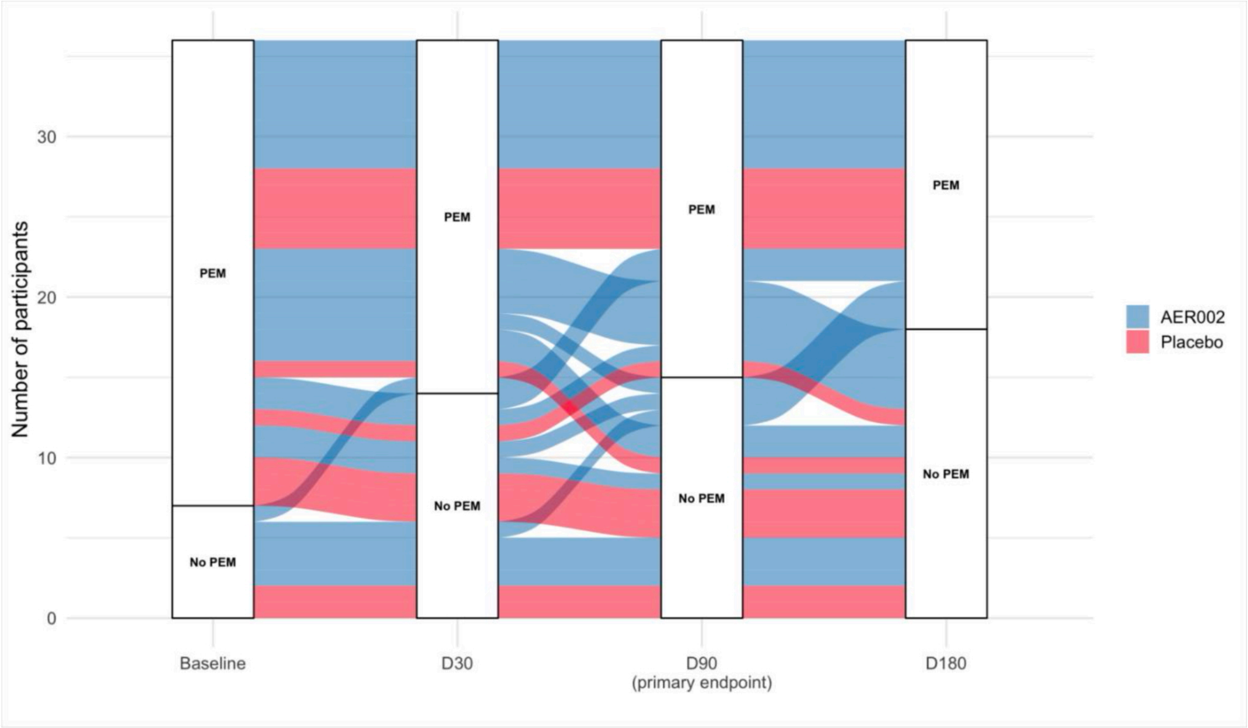


**Supplemental Figure 1. Post-exertional malaise (PEM) trends.**

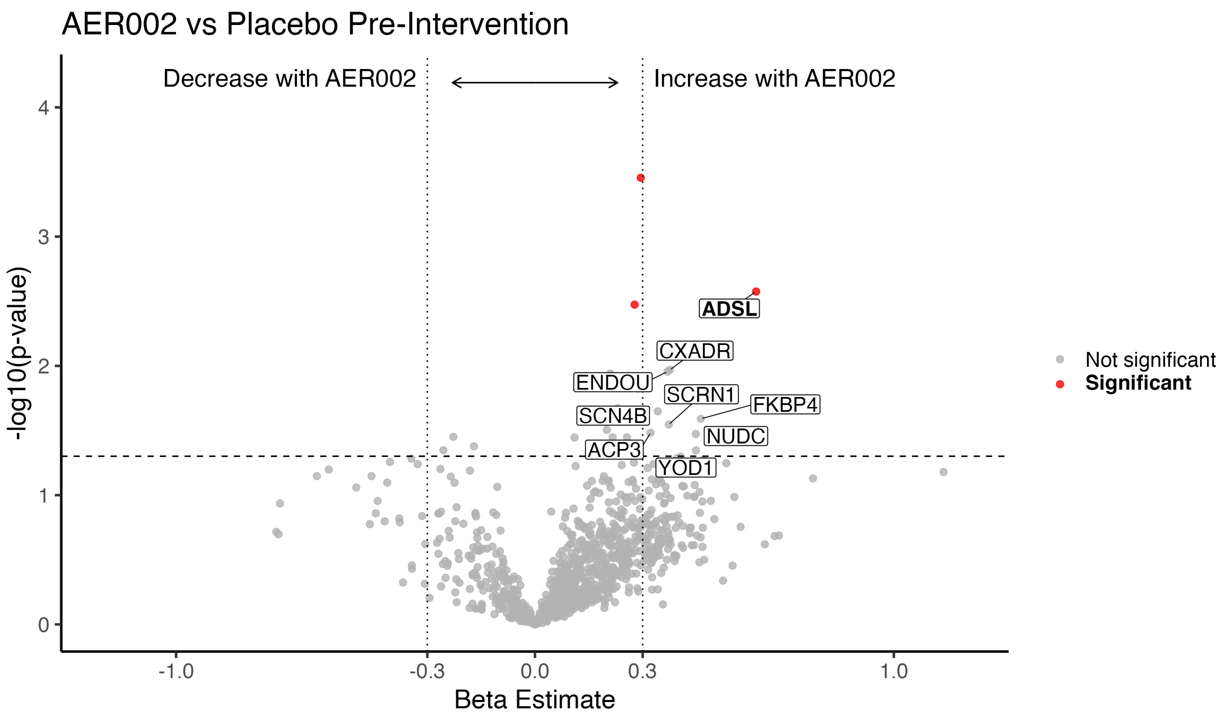


**Supplemental Figure 2. Plasma proteomics.** AER002 vs Placebo Pre-Intervention. Significant proteins (red, bolded) have Q values <0.05. Horizontal dashed line indicates unadjusted p values <0.05. Vertical dashed lines indicate beta estimates > 0.3 or <-0.3.

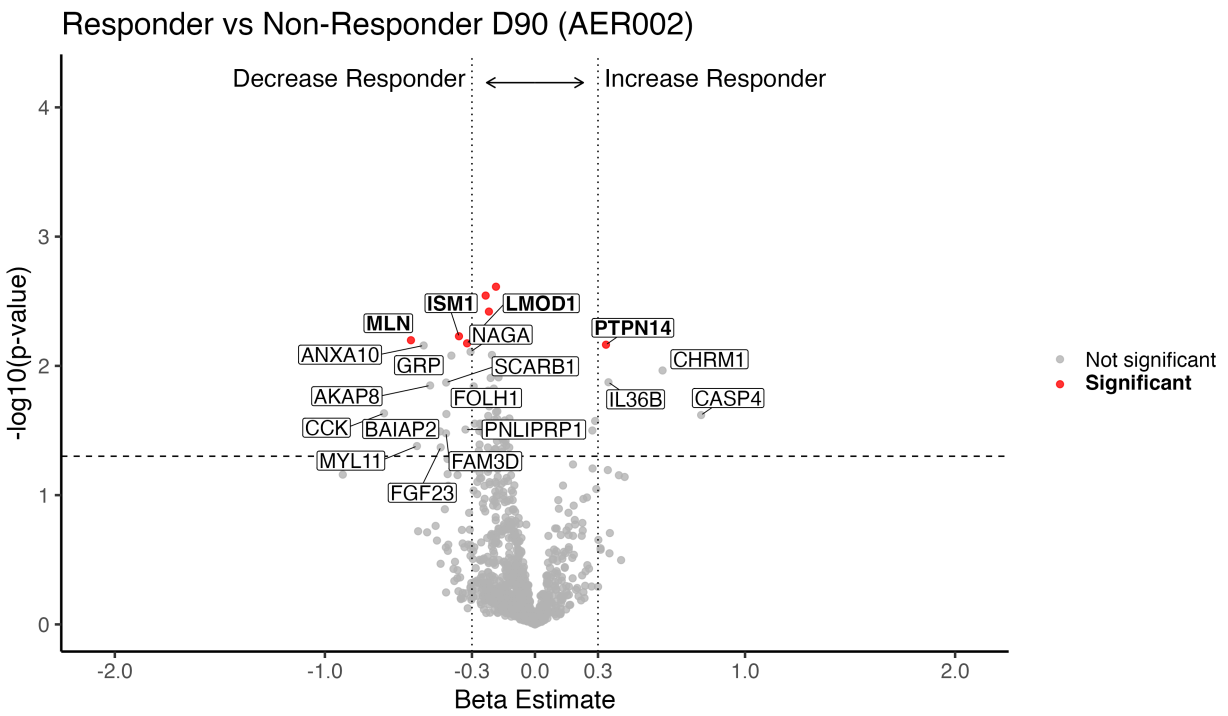


**Supplemental Figure 3. Plasma proteomics.** Responder vs Non-Responder D90 (AER002). Significant proteins (red, bolded) have Q values <0.05. Horizontal dashed line indicates unadjusted p values <0.05. Vertical dashed lines indicate beta estimates > 0.3 or <-0.3.

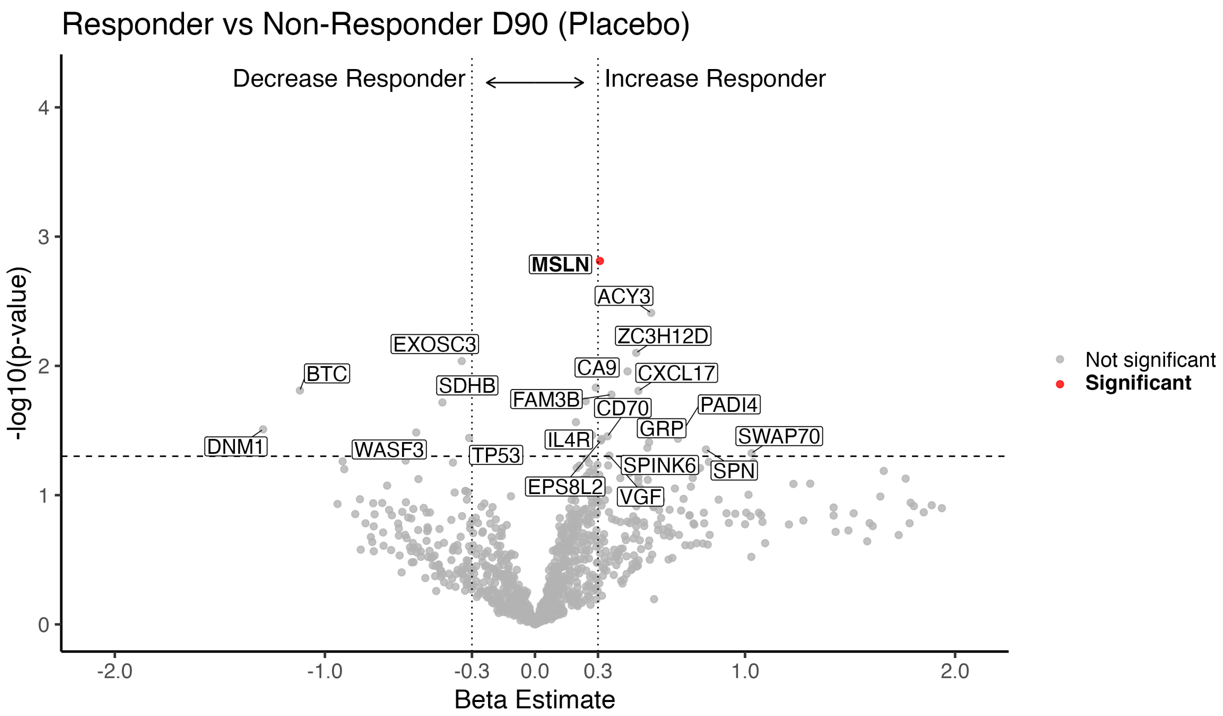


**Supplemental Figure 4. Plasma proteomics.** Responder vs Non-Responder D90 (Placebo). Significant proteins (red, bolded) have Q values <0.05. Horizontal dashed line indicates unadjusted p values <0.05. Vertical dashed lines indicate beta estimates > 0.3 or <-0.3.
